# Atrial Fibrillation/Flutter in China with Regional Disparities: Epidemiological Trends and Projections to 2050 from the Global Burden of Disease Study (1990-2021)

**DOI:** 10.64898/2026.02.02.26345382

**Authors:** Guang Li, Sijin Li, Shuang Chen, Xiancheng Xu, Weijie Wu, Canlin Li, Yuntao Tian, Laiyan Xiong, Hongsheng Liang, Heng Li

## Abstract

**Background:** Atrial fibrillation and flutter (AF/AFL) represent a major global public health challenge, contributing significantly to stroke, heart failure, and cardiovascular mortality. While previous studies have documented a rising AF/AFL burden, comprehensive comparisons of long-term trends and forecasts across regions—particularly benchmarking China against Southeast Asia, Europe, and global averages—remain limited. This study aims to quantify the AF/AFL burden across these regions from 1990 to 2021 and project trends to 2050.

**Methods:** Using data from the Global Burden of Disease Study 2021, we analysed the burden of AF/AFL from 1990 to 2021 in China, Southeast Asia, Europe, and globally. We examined incidence, prevalence, mortality, and disability-adjusted life years (DALYs). Advanced analytical methods, including Joinpoint regression, age-period-cohort modelling, decomposition analysis, Frontier analysis and Bayesian forecasting were employed to assess trends, drivers, and projections to 2050.

**Finding:** From 1990 to 2021, China experienced the most rapid increase in age-standardized incidence rate (ASIR) globally (AAPC +0.16%), with incident cases rising to 916,180, accounting for 20.43% of the global total. In contrast, Europe saw a slight decline in ASIR, while the global ASIR remained stable. China also recorded the largest increase in age-standardized prevalence rate (ASPR), whereas Europe’s ASPR declined. Despite rising incidence, China achieved the sharpest reduction in age-standardized mortality rate (ASMR; AAPC −0.45%), while Southeast Asia’s ASMR increased (AAPC +0.76%), and Europe maintained the highest ASMR globally. Frontier analysis highlighted China’s rapid efficiency improvements in mortality reduction relative to its SDI, outperforming several high-income European countries. Projections to 2050 suggest China’s ASIR and ASPR will continue to rise, whereas Europe’s are forecast to decline. Southeast Asia faces persistently increasing mortality, and global aggregates mask significant regional heterogeneity.

**Conclusion:** AF/AFL burdens are increasingly driven by population aging and metabolic risks, with heterogeneous mortality trends reflecting regional disparities in healthcare access and prevention. China’ s success in reducing mortality despite rising incidence highlights the impact of improved anticoagulation and stroke prevention, yet unchecked prevalence growth signals future complications. Southeast Asia’ s rising mortality underscores urgent needs for equitable resource allocation, while Europe’s stagnant burden reflects challenges in aging populations. Globally, prioritising primordial prevention—such as metabolic risk control—alongside targeted screening and gender-specific interventions, is critical to mitigating AF/AFL-related morbidity and mortality. Future efforts should integrate digital health technologies and address structural barriers to optimize care efficiency worldwide.

**Research in Context:** *Evidence before this study:* Prior to undertaking this analysis, we systematically reviewed the existing epidemiological literature on atrial fibrillation and atrial flutter (AF/AFL), with a particular emphasis on global and regional comparative studies. Our searches covered PubMed, Embase, Web of Science, the Cochrane Library, and the Global Burden of Disease (GBD) repository from January 1990 to December 2023, without language restrictions. Key terms included “atrial fibrillation,” “atrial flutter,” “global burden,” “epidemiology,” “trend,” and “GBD.” We included studies providing representative estimates of AF/AFL burden and excluded small-sample or non-age-standardized reports. Previous analyses indicated a rising global AF/AFL burden, largely due to population aging and improved detection. However, comprehensive assessments capturing temporal dynamics, risk drivers, and forecasting across major world regions—especially benchmarking China, Southeast Asia, and Europe against global patterns—remained limited. Most studies focused on isolated regions or short spans, lacking integrative multidimensional approaches such as age-period-cohort modeling, decomposition, or Bayesian forecasting.

*Added value of this study:* This study provides a comprehensive and comparative assessment of the atrial fibrillation and atrial flutter (AF/AFL) burden across China, Southeast Asia, Europe, and globally from 1990 to 2021, utilizing the latest GBD 2021 data and advanced statistical methodologies, including Joinpoint regression, age-period-cohort modeling, Bayesian forecasting, decomposition analysis, and data envelopment frontier analysis. Our analysis reveals significant regional disparities against a backdrop of global stability: while the global age-standardized incidence rate (ASIR) remained stable (52·51 in 1990 vs. 52·12 in 2021), China experienced the most rapid increase worldwide (ASIR rising from 42·63 to 44·92), with a substantial number of new cases (916,180), accounting for 20·43% of the global total (4,484,926 cases). In contrast, Europe recorded a slight decline in ASIR. China also demonstrated the most pronounced increase in prevalence globally, while Europe’s age-standardized prevalence rate (ASPR) declined and the global rate remained largely unchanged. Notably, China achieved a significant reduction in mortality (age-standardized mortality rate [ASMR] declining from 4·93 to 4·33) despite rising incidence, sharply contrasting with Southeast Asia, where ASMR increased from 2·94 to 4·06 (estimated annual percentage change +1·07%)—trends potentially associated with structural challenges in Southeast Asia—while Europe maintained the highest ASMR globally (5·10 in 2021) despite interventions. We further identified key drivers: population growth and aging accounted for the majority of the case increase in China, consistent with global demographic trends, while metabolic risk factors accelerated this trend. Gender and age analyses revealed a global pattern of later-life predominance in women and earlier onset in middle-aged groups, particularly pronounced in China. Our projections to 2050 indicate a continued rise in ASIR and ASPR in China, reinforcing its significant and growing contribution to the global AF/AFL burden, whereas other regions face divergent challenges—Southeast Asia is projected to experience persistently increasing mortality pressure, while Europe must address persistently high disability-adjusted life year (DALY) rates, masking mortality gains in an aging population.

*Implications of all the available evidence:* The collective evidence from this study and previous research underscores that AF/AFL remains a critical and growing public health challenge worldwide, characterized by heterogeneous patterns across regions when viewed against the global aggregate. China’s success in reducing mortality within a rising incidence environment highlights the potential of enhanced clinical management and stroke prevention, yet its unchecked prevalence growth—especially among younger cohorts—signals a looming surge in complications absent strengthened primary prevention, a concern mirrored in many developing economies. Southeast Asia’s rising mortality underscores urgent needs for improved access to anticoagulation and rhythm control, while Europe’s stagnant burden reflects challenges in managing an aging population efficiently. These findings advocate for regionally tailored strategies that align with global frameworks but address local disparities—integrating primordial prevention (e.g., metabolic risk control) with early detection, gender-specific treatment, and equitable resource allocation. Future research should prioritize mechanistic studies of AF/AFL subtypes, real-world intervention assessments, and the integration of digital health technologies for scalable screening and management, thereby informing coordinated global actions to mitigate the evolving burden of AF/AFL.

## Introduction

Atrial fibrillation/flutter (AF/AFL) is recognized as a prevalent cardiac arrhythmia that poses a significant risk for various complications, including stroke and heart failure, leading to substantial morbidity and mortality rates among affected individuals.^1^ The economic implications of AF/AFL are considerable, as it is associated with increased hospitalization rates and ongoing medical management expenses.^2,3^ This not only imposes a substantial burden on families but also significantly compromises patients’ quality of life through functional impairment and psychosocial distress. Moreover, systemic challenges, such as the suboptimal allocation of healthcare resources for procedures like catheter ablation and the absence of personalized preventive management, continue to impede optimal disease control.. Consequently, there is a pressing need for research aimed at elucidating the burden of AF/AFL and informing public health initiatives to improve patient outcomes.^4^

Current literature highlights considerable variability in the epidemiological indicators of AF/AFL across different populations.^5^ While previous studies have documented trends in AF/AFL prevalence.^6^ comprehensive assessments capturing the long-term epidemiological trends, regional disparities, and drivers of AF/AFL burden across diverse global regions—particularly comparing regions at different stages of development and healthcare system maturity—remain limited. Understanding these trends is crucial for recognizing the broader impact of AF/AFL and for developing informed healthcare policies and resource allocation strategies. China, the most populous nation worldwide, exemplifies this conundrum: its rapid economic growth has been coupled with vigorous health-system reforms and noticeable improvements in the identification and management of AF/AFL. However, the country is concurrently facing a rising tide of metabolic risk factors and a significant aging population.^7,8^ Indeed, robust longitudinal data that situate China’s AF/AFL trajectory within a global comparative framework remain markedly limited. This scarcity is particularly evident in comparisons with regions such as Southeast Asia—which shares analogous demographic transitions but contends with distinct healthcare access barriers—or Europe, where advanced health systems coexist with persistent challenges related to aging populations and cost inefficiencies. Moreover, while global aggregates offer broad benchmarks, they often obscure profound underlying disparities in healthcare infrastructure, data quality, risk factor distribution, and resource allocation. Collectively, there exists a dearth of comparative data that positions China alongside Southeast Asia, the relatively advanced health systems of Europe, and the global average over the past three decades.^9^

To address these gaps, this research employs advanced statistical methodologies, including age-standardized rates and temporal trend analysis, utilizing techniques such as Joinpoint regression and Age-Period-Cohort (APC) models. These methodologies are designed to enhance the accuracy of trend identification and to facilitate a nuanced understanding of the disease burden across different demographics. By integrating diverse data sources and employing robust analytical frameworks, the study aims to quantify the variations in AF/AFL burden across distinct geographical regions and to identify the underlying factors contributing to these differences.

The primary objective of this study is to provide a comprehensive assessment of the burden of AF/AFL, focusing on key epidemiological indicators such as incidence, prevalence, mortality rates, and disability-adjusted life years (DALYs). By dissecting these indicators, the research aspires to illuminate the multifaceted nature of AF/AFL and its implications for public health. Furthermore, the findings are expected to serve as an invaluable resource for policymakers and healthcare providers, facilitating tailored interventions and strategic planning that address the growing burden of AF/AFL.

In summary, this research endeavors to fill the existing knowledge gaps regarding the burden of AF/AFL by employing sophisticated statistical approaches to analyze a rich dataset spanning multiple countries and years. The insights garnered from this study will not only advance our understanding of AF/AFL trends but will also contribute to the development of targeted public health strategies aimed at mitigating the impact of this prevalent arrhythmia on global health outcomes. This study is poised to significantly influence discourse on AF/AFL-related policy formulation, decision evaluation, resource allocation, and regulation within healthcare systems worldwide.

## Materials and Methods

### Data source

We obtained data from the Global Burden of Disease Study (GBD 2021) database on February 25, 2025, which is an updated version of the GBD 2019 project, providing extensive epidemiological information for 204 countries and regions from 1990 to 2021.^10^ The GBD system collects data from various sources, including national population censuses, household surveys, civil registration systems, vital statistics, disease registration, health service records, and other relevant notifications.^11^ GBD 2021 employs a suite of advanced statistical models to ensure comprehensive and comparable estimates across diseases, injuries, and risk factors. Key models include: DisMod-MR 2·1, the Cause of Death Ensemble model (CODEm), Meta-Regression Bayesian Regularized Trimmed (MR-BRT), Spatiotemporal Gaussian Process Regression (ST-GPR), and the Bayesian Age-Period-Cohort (BAPC) model. It is important to note that The GBD study adheres to strict ethical standards during its original data aggregation and processing stages. Its data sources include publicly available national censuses, civil registration and vital statistics systems, scientific literature, and disease surveillance records from various regions. All personal identifiers are removed before the data enter the GBD modeling pipeline, and researchers only have access to aggregated statistical results that cannot be traced back to any specific individual. For our purposes, we accessed the data through the query tool provided at the link, from which we extracted specific causes of death, various observation indicators, stratified by age, gender, year, and region. The GBD study protocol has been approved by the Institutional Review Board (IRB) of the University of Washington. According to the Declaration of Helsinkiand internationally recognized ethical guidelines (such as the CIOMS guidelines), such studies are generally eligible for exemption from ethical review. In accordance with journal policy, the analysis code used in this study is available from the corresponding author upon reasonable request. It is worth noting that the data analyzed in this study pertains to individuals aged 30 and above, consistent with the scope of the GBD 2021 report, including information from China, Southeast Asia, Europe, and global populations.

### Disease burden description

In this research, we quantified the burden of AF/AFL by examining several epidemiological indicators, including the annual incidence, prevalence, mortality, and DALYs, along with their respective age-standardized rates (ASRs) per 100,000 people. To facilitate meaningful comparison across different populations, we calculated the ASRs based on the World Health Organization’s (WHO) 2000-2025 standard population. All estimates are presented alongside 95% CI, corresponding to the 2·5th and 97·5th percentiles derived from 100 uncertainty distributions. Utilizing Age-Standardized Incidence Rate (ASIR), Age-Standardized Prevalence Rate (ASPR), and Age-Standardized Mortality Rate (ASMR) enables accurate comparisons between populations with varying age structures and sizes, thereby enhancing analytical precision. The scope of our analysis encompasses the burden of AF/AFL in China, Europe, Southeast Asia, and globally. Additionally, we examined how these metrics distribute across different age groups and genders, analyzing the multifaceted impact of the disease burden from multiple perspectives.

### Trends analysis

In order to examine the temporal trends associated with the disease burden of AF/AFL from 1990 to 2021, we utilized the Joinpoint regression model. This statistical method is well-regarded in epidemiology for its effectiveness in analyzing time-series data and identifying significant trend changes. The model determines residuals through least squares, aiming to reduce discrepancies between actual and projected values, effectively pinpointing inflection points without depending on subjective criteri.^12^ All trend analyses were performed on age-standardized rates (ASRs), including the Age-Standardized Incidence Rate (ASIR), Age-Standardized Prevalence Rate (ASPR), Age-Standardized Mortality Rate (ASMR), and Age-Standardized DALY Rate (ASDR), each expressed per 100,000 person or years. Furthermore, to assess the evolving trends, we calculated both the Average Annual Percentage Change (AAPC) and the Annual Percentage Change (APC) for each identified segment. The AAPC and APC represent relative changes (percentage change per year) in these ASRs. An AAPC or APC value exceeding zero indicated a rising trend, while values below zero implied a declining trend; a value of zero was interpreted as a stable trend. We established a threshold of P < 0·05 to denote statistical significance.

### Age-Period-Cohort (APC) model

For comprehensive evaluation, we applied the APC model, a robust statistical framework that enables researchers to analyze the independent and combined effects of age, period, and birth cohort on health outcomes and population trends. Period effects are caused by changes in social, economic, cultural, or physical environments, affecting all age groups equally, and differences within the same birth year group can also be attributed to cohort effects. Period and cohort relative risk (RR) refers to the age-specific ratio of each period and cohort relative to the control group. Specifically, we used this model to analyze the prevalence, incidence rate, mortality rate, and DALYs of AF/AFL patients in the observation area of our article. Analyses were performed using the web-based Age-Period-Cohort Analysis Tool developed by the Division of Cancer Epidemiology and Genetics at the US National Cancer Institute (available at: [https://analysistools.cancer.gov/apc/]). We obtained estimated parameters (including Period RR, Cohort RR, and Net Drift) from this tool. This approach allows us to better understand the trends of atrial fibrillation/atrial fibrillation-like episodes, providing valuable insights into this critical public health issue.^13,14^

### Bayesian Age-Period-Cohort (BAPC) model

We use the BAPC model to predict the disease burden of AF/AFL in the future. The model is selected because it can effectively clarify the age, period and cohort effects, and can capture nonlinear trends and group specific risk profiles.^15^ The model employs the Integrated Nested Laplacian Approximation (INLA) algorithm for Bayesian inference. To account for the temporal structure inherent in age, period, and cohort effects, we utilized second-order random walk (RW2) priors for each temporal dimension. This choice of prior distribution imposes smoothness constraints, penalizing abrupt changes and thereby providing robust estimations of the temporal trends. Hyperparameters for the precision (inverse variance) of the RW2 priors were assigned Gamma hyperpriors (shape = 1, rate = 0·00005), following default recommendations within the BAPC package and established practices for stabilizing variance estimation in Bayesian APC models. The integrated nested Laplacian approximation (INLA) is applied to improve the accuracy, which surpasses the traditional linear method. Positive BAPC values above zero indicate an increasing trend, while negative values indicate a decreasing trend. Therefore, based on GBD 2021 research data, this study uses this model to predict the trend ofASPR, ASIR, ASMR and ASDR ofpatients in the observed region from 2022 to 2050.

### Decomposition analysis

In the research of GBD, decomposition analysis is often used to quantify the impact of population and risk factors. The change of disease burden is attributed to population aging, population growth and epidemiological changes, so as to reveal the potential factors leading to the observed changes.^16^ The purpose of this study is to explore the driving force behind the change of disease burden by using decomposition analysis method. All the analysis and visualization in this study were performed in R language (version 4·3·1). When the bilateral P<0·05, the difference was considered statistically significant.

### Frontier analysis

In order to explore and horizontally compare the prevention and treatment of disease burden in more dimensions, we also use the data envelopment analysis (DEA) method based on linear programming for frontier analysis. DEA determines the best performance by constructing a “boundary” containing all decision-making units, and the units below the boundary are considered invalid. In order to evaluate the robustness of DEA, we use a bootstrap process with 100 iterations to generate multiple resampled datasets to evaluate the stability. Local weighted regression (LOESS) was used to visualize the final results to illustrate the trends and performance of ASMR and ASDR attributed to AF/AFL in the observed region from 1990 to 2021.^17^

### The Socio-demographic Index (SDI)

The Socio-demographic Index (SDI) is a composite metric developed by the Institute for Health Metrics and Evaluation (IHME) to quantify the development level of regions or countries. It integrates three core dimensions: (1) lag-distributed income (LDI) pe rcapita, adjusted for purchasing power parity (PPP), which serves as an indicator of economic resources and is sourced from the World Bank Development Indicators; (2) average educational attainment, measured by the mean years of schooling among individuals aged 15 years and older, derived from UNESCO databases and national censuses; and (3) fertility rate, represented by the total fertility rate (TFR) for women under age 25, based on data from the UN Population Division. Each of these components is first normalized to a scale of 0 to 100, where 0 represents the minimum observed value globally from 1990 to 2021 and 100 represents the maximum value. The SDI value is then computed as the geometric mean of these three normalized components, resulting in a score ranging from 0 (lowest development) to 1 (highest development). To facilitate comparative analyses, regions are categorized into quintiles based on their SDI values: low (0–0·20), low-middle (0·21–0·40), middle (0·41–0·60), high-middle (0·61–0·80), and high (0·81–1·00). This index is particularly valuable for examining health system efficiency and epidemiological patterns across different development contexts, as it enables robust cross-regional comparisons of disease burden and resource allocation efficiency, with detailed methodology and global estimates provided by the GBD Collaborative Network (2021).

### Reporting Guidelines

The reporting of this observational study follows the Strengthening the Reporting of Observational Studies in Epidemiology (STROBE) guidelines.

## Results

### Overall burden of AF/AFL from 1990 to 2021

Figure 1 provides an overview of the workflow and key emphases of this study. Between 1990 and 2021 the absolute number of incident AF/AFL cases in China rose from 306 585 (95% CI: 234 243-404 868) to 916 180 (95% CI: 707 384-1 201 381). ASIR increased modestly, from 42·63 to 44·92 per 100 000 (AAPC 0·155 %, 0·051-0·260), the steepest rise among the four macro-regions examined. Southeast Asia followed a similar upward trajectory (1990: 318 971 cases, ASIR 52·28; 2021: 900 234 cases, ASIR 53·02), whereas Europe (1990: 645 411, 59·47; 2021: 957 812, 58·93) and the globe (1990: 2·01 million, 52·51; 2021: 4·33 million, 52·12) experienced slight net declines in ASIR (Europe AAP=0·035 % verse Global AAPC=0·027 %). These divergent slopes highlight a significant increase in ASIR within China, which may reflect advancements in detection technologies alongside the heterogeneous pace of preventive cardiology development across regions.

**Figure 1.**
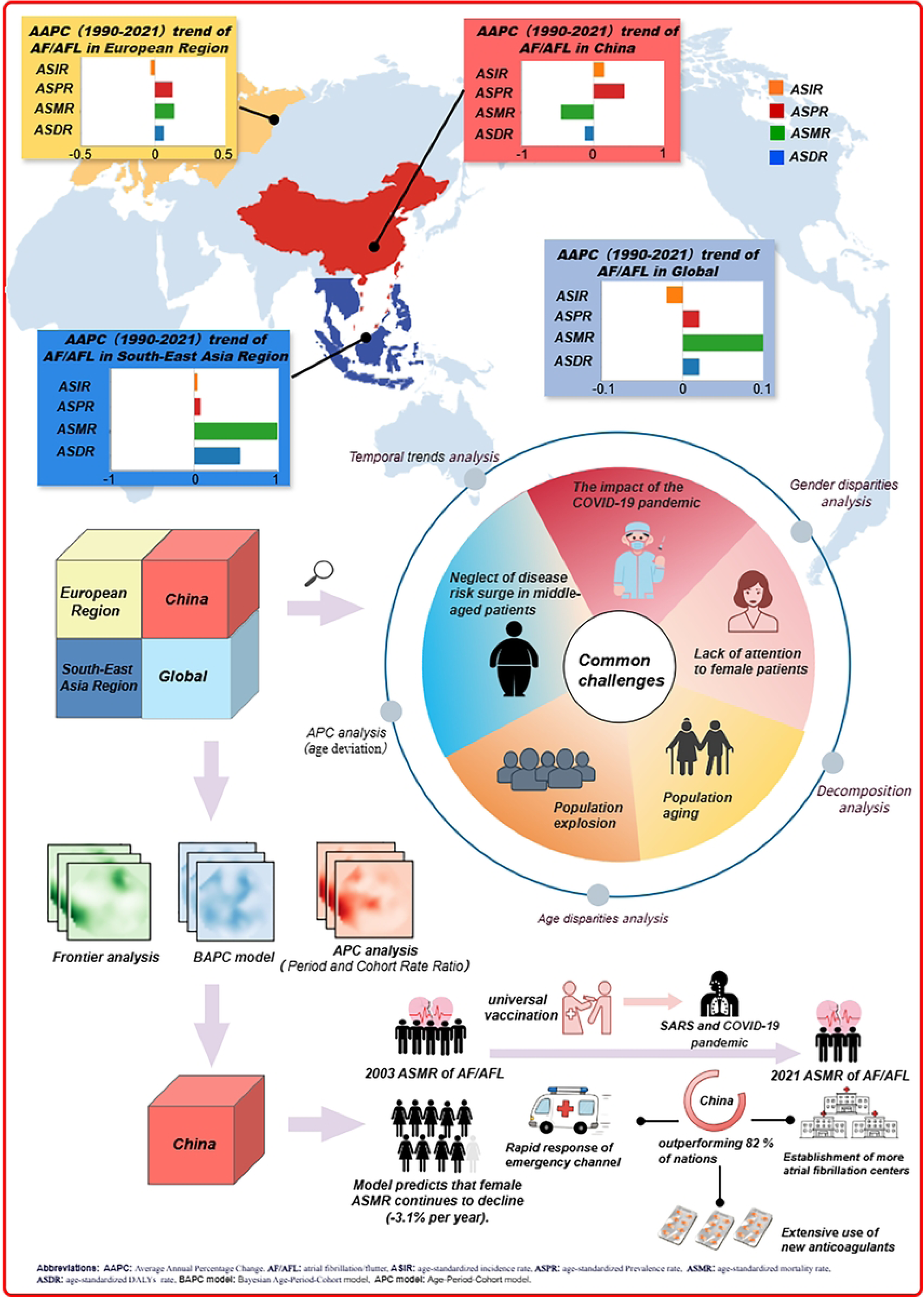
Central illustration and flow chart of the study design. Trends in Age - Standardized Rates of Atrial Fibrillation/Flutter (AF/AFL) From 1990 to 2021, Analytical Frameworks, Shared Challenges, and China’s Response.

Prevalent cases in China expanded almost 3·4-fold, from 3·20 million (2·52-4·17) to 10·78 million (8·53-14·01). ASPR climbed from 457·7 to 524·0 per 100 000 (AAPC 0·433 %, 0·324-0·543), again the largest increment worldwide. Southeast Asia’s ASPR edged upward from 549·4 to 562·4, Europe’s fell from 713·0 to 562·4, and the global rate inched from 616·6 to 620·5. The sustained growth in China reflects both demographic momentum and an apparent epidemiological shift-possibly driven by hypertension control gaps, dietary sodium excess and rising obesity-that outweighs improvements in secondary prevention.

Deaths attributable to AF/AFL in China quadrupled from 16 449 (13 240-20 521) to 64 728 (51 765-77 729), yet age-standardized mortality rate (ASMR) fell from 4·93 to 4·33 per 100 000 (AAPC) 0·45 %), the sharpest decline among all studied regions. Southeast Asia conversely saw ASMR rise from 2·94 to 4·06 (AAPC 0·76 %), Europe from 4·92 to 5·10, and the globe from 4·24 to 4·36. These patterns suggest that China’s health system has been comparatively effective at averting premature deaths once AF/AFL is manifest, whereas other regions - especially Southeast Asia - struggle to deliver guideline-concordant stroke prophylaxis or rate/rhythm control.

DALYs in China surged from 508 610 (395 853-638 618) to 1·65 million (1·30-2·06), but age-standardized DALY rate (ASDR) slipped slightly from 93·29 to 89·76 per 100 000 (AAPC - 0·12%). Southeast Asia, Europe and the global average all recorded net increases in ASDR. The combination of rising absolute DALYs but falling rates in China underscores rapid population ageing: years lived with disability accumulate, yet the average individual loses fewer life-years than three decades ago (Table 1, 2).

**Table 1.**
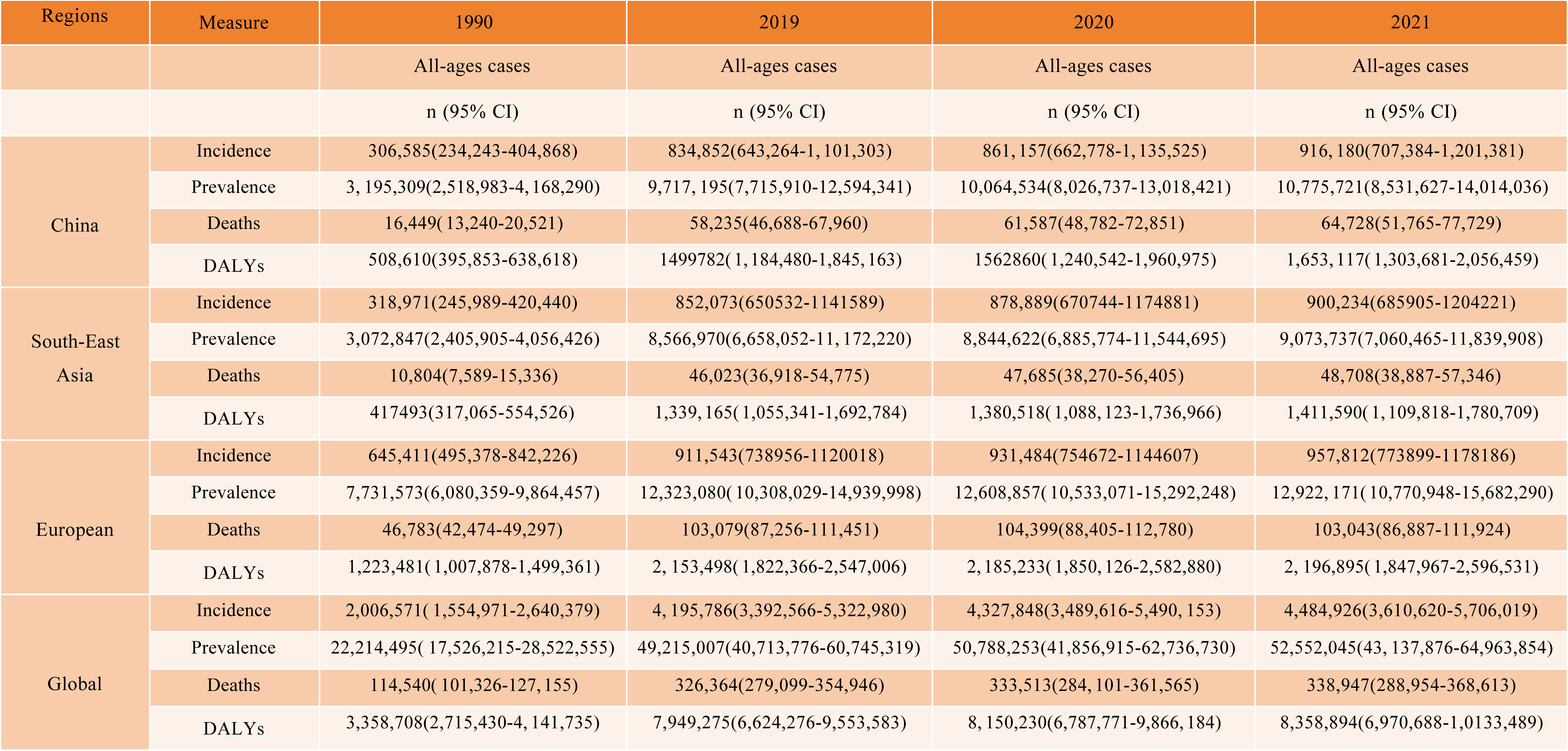
All-age cases incidence, prevalence, mortality, and DALYs rates ofAF/AFL in China, South-East Asia Region, European Region and Global in 1990, 1991,2020 and 2021.

**Table 2.** Age-standardized incidence, prevalence, mortality, DALYs rates and corresponding AAPC of RHD in China, South-East Asia Region, European Region and Global in 1990, 1991,2020 and 2021.

| Regions | Measure | 1990 | 2019 | 2020 | 2021 | 2019-2021 AAPC |
| --- | --- | --- | --- | --- | --- | --- |
|  |  | Age-standardized rates per 100,000 | Age-standardized rates per 100,000 | Age-standardized rates per 100,000 | Age-standardized rates per 100,000 | AAPC |
|  |  | n (95% CI) | n (95% CI) | n (95% CI) | n (95% CI) | n (95% CI) |
| China | Incidence | 42.628(32.4-56.463) | 43.744(34.002-57.952) | 43.745(34.074-57.836) | 44.923(34.959-59.421) | 0.1554(0.0514-0.2596) |
|  | Prevalence | 457.72(358.933-594.962) | 506.881(404.189-656.054) | 507.436(405.57-656.48) | 524.004(418.147-681.226) | 0.4331(0.3237-0.5426) |
|  | Deaths | 4.933(3.882-6.166) | 4.3(3.432-5.053) | 4.329(3.378-5.164) | 4.327(3.426-5.227) | -0.4540(-0.7833--0.1237) |
|  | DALYs | 93.285(75.142-115.499) | 88.278(71.235-108.053) | 88.537 (72.263-110.945) | 89.757(72.135-109.667) | -0.1235(-0.3158-0.0691) |
| South-East Asia Region | Incidence | 52.275(39.473-69.415) | 53.196(40.192-70.787) | 53.213(40.175-70.659) | 53.022(40.03-70.511) | 0.0458(0.0358-0.0557) |
|  | Prevalence | 549.407(428.113-715.539) | 563.664(439.636-733.856) | 563.793(439.742-735.036) | 562.436(439.228-732.191) | 0.0765(0.0717-0.0814) |
|  | Deaths | 2.937(2.11-4.139) | 4.087(3.265-4.869) | 4.084(3.276-4.814) | 4.057 (3.236-4.761) | 1.0729( 0.7596-1.3871) |
|  | DALYs | 80.358(61.518-105.392) | 95.213(75.709-118.381) | 95.031(75.963-118.01) | 94.513(75.574-117.078) | 0.5508(0.3841-0.7177) |
| European | Incidence | 59.472(46.039-76.833) | 57.63(47.053-70.598) | 58.057(47.42-70.921) | 58.925(47.79-72.106) | -0.035(-0.0484--0.0216) |
|  | Prevalence | 713.031(563.465-907.355) | 726.543(609.787-873.622) | 732.795(614.961-880.832) | 741.425(619.282-890.718) | 0.1261(0.1096-0.1425) |
|  | Deaths | 4.917(4.403-5.216) | 5.282(4.497-5.697) | 5.237(4.465-5.645) | 5.098(4.33-5.522) | 0.1323(-0.0352-0.3001) |
|  | DALYs | 117.113(97.255-142.263) | 120.663( 101.673-143.634) | 120.385( 101.312-143.388) | 119.672(99.9-142.829) | 0.0586(0.0177-0.0995) |
| Global | Incidence | 52.51(40.385-69.006) | 51.265(41.338-64.954) | 51.515(41.429-65.285) | 52.116(41.854-66.231) | -0.0271(-0.0345--0.0197) |
|  | Prevalence | 616.576(485.217-795.265) | 611.753(506.887-754.957) | 614.423(508.074-758.902) | 620.507(511.357-768.875) | 0.019(0.0038-0.0343) |
|  | Deaths | 4.239(3.694-4.705) | 4.449(3.781-4.846) | 4.406(3.73-4.788) | 4.363(3.691-4.752) | 0.1061(0.0309-0.1814) |
|  | DALYs | 100.815(82.825-122.617) | 101.728(85.228-121.828) | 101.378(84.797-122.091) | 101.398(84.89-122.412) | 0.022(-0.0336-0.0777) |

### Temporal trends in AF/AFL burden from 1990 to 2021

Joinpoint regression identified two distinct epochs in China: (i) 2000-2005 (APC 1·60 %) and (ii) 2015-2021 (APC 1·21 %), both statistically significant (p < 0·05) (Fig. 2A). A transient downturn (2005-2010) coincided with nationwide antihypertensive campaigns; however, the long-term slope remains upward, mirroring Southeast Asia and contrasting with Europe’s steady decline (Fig. 2B and 2C). After 2019, the COVID-19 pandemic coincided with an abrupt spike in ASIR in China, Europe and globally - plausibly through delayed diagnoses, inflammatory milieu and healthcare disruptions—whereas Southeast Asia recorded no inflection.

**Figure 2.**
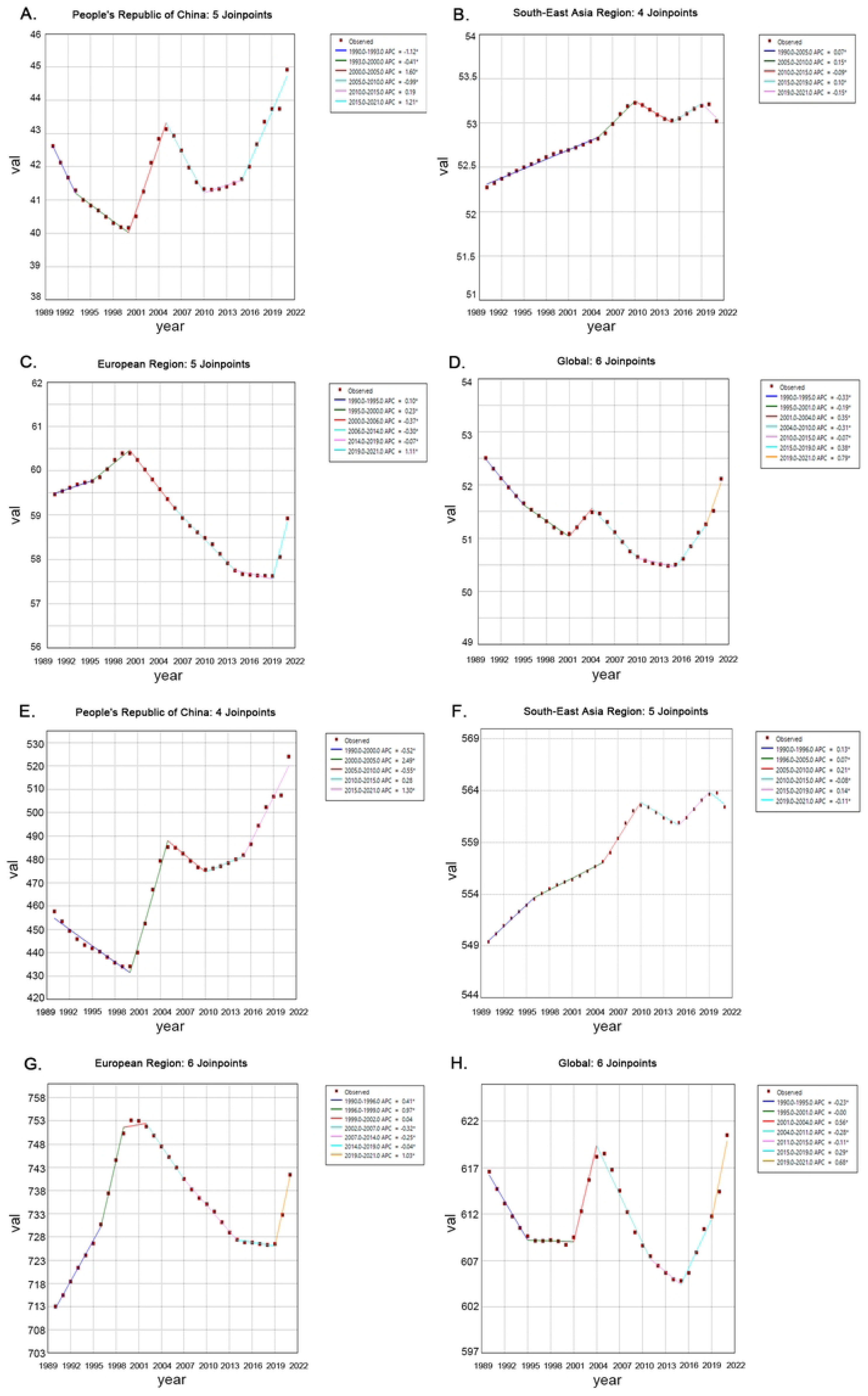
EAPC trends of ASIR and ASPR for AF/AFL. EAPC (Estimated Annual Percentage Change) trends of ASIR (Age-Standardized Incidence Rate) and ASPR (Age-Standardized Prevalence Rate) for AF/AFL in China, South-East Asia, Europe and Global in1990-2021 (all graphs share identical axes: X-axis = year [1990–2021]; Y-axis = age-standardized metric value; dark red dots =annual observed values): (A) China (ASIR): Segments: dark blue (1990-1993), green (1993-2000), red (2000-2005), light blue (2005-2010), purple (2010-2015), blue (2015-2021). Joinpoints: 5. (B) South-East Asia (ASIR): Segments: dark blue (1990-2005), green (2005-2010), red (2010-2015), light blue (2015-2019), purple (2019-2021). Joinpoints: 4. (C) Europe (ASIR): Segments: dark blue (1990-1995), green (1995-2000), red (2000-2006), light blue (2006-2014), purple (2014-2019), blue (2019-2021). Joinpoints: 5. (D) Global (ASIR).Segments: dark blue (1990-1995), green (1995-2001), red (2001-2004), light blue (2004-2010), purple (2010-2015), blue (2015-2019), orange (2019-2021). Joinpoints: 6. (E) China (ASPR) Segments: dark blue (1990-2000), green (2000-2005), red (2005-2010), light blue (2010-2015), purple (2015-2021). Joinpoints: 4. (F) South-East Asia (ASPR) Segments: dark blue (1990-1996), green (1996-2005), red (2005-2010), light blue (2010-2015), purple (2015-2019), blue (2019-2021). Joinpoints: 5. (G) Europe (ASPR): Segments: dark blue (1990-1996), green (1996-1999), red (1999-2002), light blue (2002-2007), purple (2007-2014), blue (2014-2019), orange (2019-2021). Joinpoints: 6. (H) Global (ASPR): Segments: dark blue (1990-1995), green (1995-2001), red (2001-2004), light blue (2004-2011), purple (2011-2015), blue (2015-2019), orange (2019-2021). Joinpoints: 6.

The ASPR of China exhibited sustained growth punctuated by a brief 2005-2010 dip (APC - 0·3 %) (Fig. 2E). The most recent segment (2015-2021, APC 1·30 %) aligns with intensified secondary prevention (oral anticoagulation uptake > 40 % among high-risk patients) but is insufficient to offset demographic expansion. Parallel trajectories in Southeast Asia diverge after 2019 (Fig. 2F), when prevalence accelerated further, while Europe plateaued (Fig. 2G).

China’s ASMR declined in two windows (2004-2007 APC) 3·78 %; 2010-2013 APC - 4·12 %), interrupted by a modest rebound (2007-2010 APC 0·67 %) (Fig. 3A). The net downward drift reflects cumulative gains in acute stroke care, anticoagulation and integrated AF management. Conversely, Southeast Asia, Europe and the globe experienced rising ASMR until 2019; thereafter Europe and the globe recorded sharp COVID-era declines, whereas China’s trajectory remained comparatively stable.

**Figure 3.**
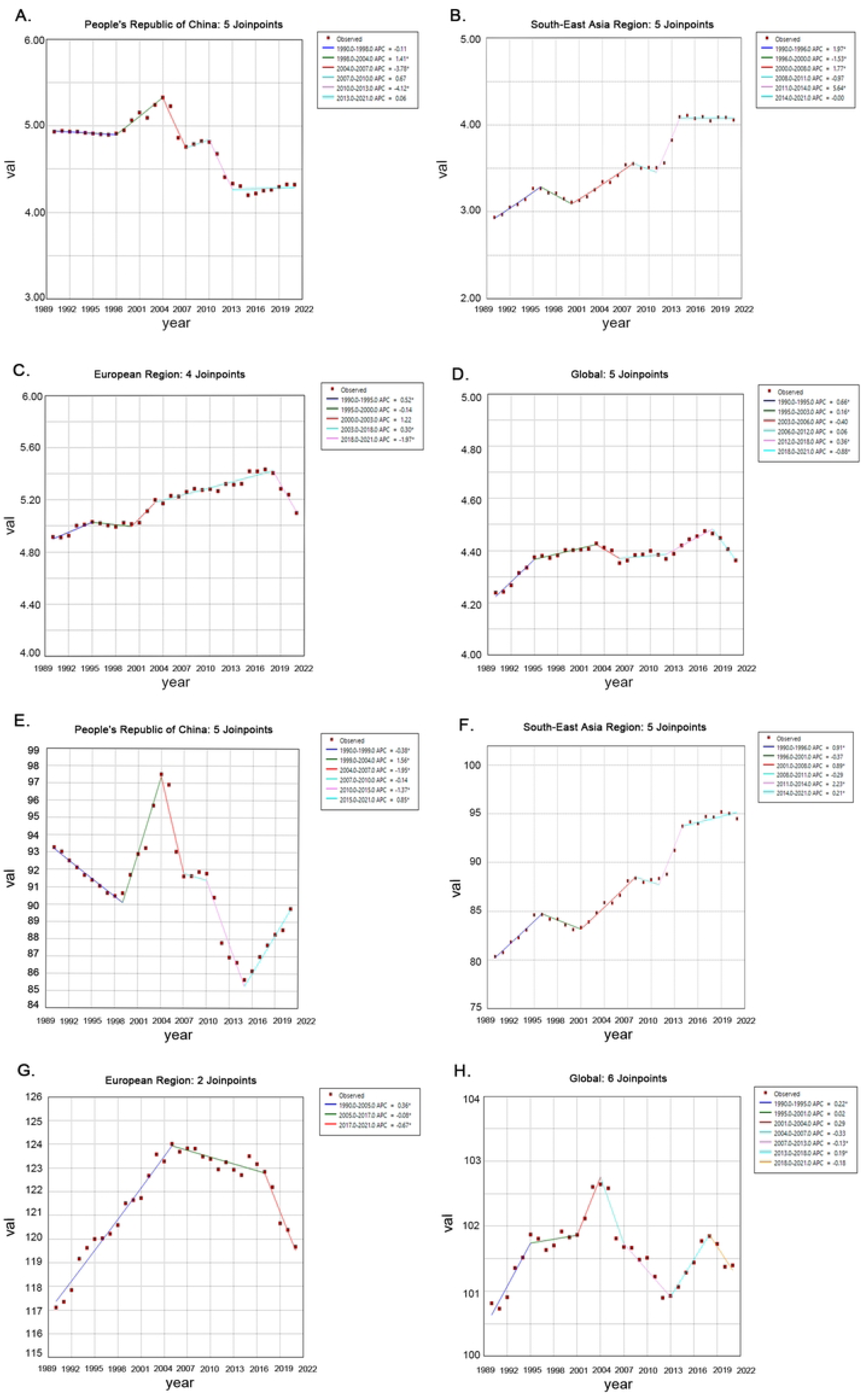
EAPC trends of ASMR and ASDR for AF/AFL. EAPC (Estimated Annual Percentage Change) trends of ASMR (Age-Standardized Mortality Rate) and ASDR (Age-Standardized DALYs Rate) for AF/AFL in 1990-2021. (all graphs share identical axes: X-axis = year [1990–2021]; Y-axis = age-standardized metric value; dark red dots =annual observed values): (A) China (ASMR): Segments: dark blue (1990-1998), green (1998-2004), red (2004-2007), light blue (2007-2010), purple (2010-2013), blue (2013-2021). Joinpoints: 5. (B) South-East Asia (ASMR): Segments: dark blue (1990-1996), green (1996-2000), red (2000-2008), light blue (2008-2011), purple (2011-2014), blue (2014-2021). Joinpoints: 5. (C) Europe (ASMR): Segments: dark blue (1990-1995), green (1995-2000), red (2000-2003), light blue (2003-2018), purple (2018-2021). Joinpoints: 4. (D) Global (ASMR): Segments: dark blue (1990-1995), green (1995-2003), red (2003-2006), light blue (2006-2012), purple (2012-2018), blue (2018-2021). Joinpoints: 5. (E) China (ASDR): Segments: dark blue (1990-1999), green (1999-2004), red (2004-2007), light blue (2007-2010), purple (2010-2015), blue (2015-2021). Joinpoints: 5. (F) South-East Asia (ASDR): Segments: dark blue (1990-1996), green (1996-2001), red (2001-2008), light blue (2008-2011), purple (2011-2014), blue (2014-2021). Joinpoints: 5. (G) Europe (ASDR). Segments: dark blue (1990-2005), green (2005-2017), red (2017-2021). Joinpoints: 2. (H) Global (ASDR): Segments: dark blue (1990-1995), green (1995-2001), red (2001-2004), light blue (2004-2007), purple (2007-2013), blue (2013-2018), orange (2018-2021). Joinpoints: 6.

China’s ASDR fell between 2004 and 2015, yet an upswing since 2016 (APC 1·56 %) suggests that quality-of-life improvements are eroding under the weight of aging, multimorbidity and healthcare inequality (Fig. 3E). Europe achieved sustained post-2004 reductions that steepened during the pandemic (Fig. 3G), whereas Southeast Asia’s ASDR rose monotonically (Fig. 3F).

### Age-specific burden profiles in AF/AFL

Across all regions, crude incidence peaks at 65-74 years. In China, however, the modal age shifted downwards from 70-74 in 1990 to 65-69 in 2021, hinting at earlier onset driven by cardiometabolic risk accumulation (Fig.4A). By contrast, Southeast Asia, Europe and the global average recorded upward shifts to 70-74 years. Notably, China is the only region where incidence in (40-49)-year-olds declined continuously after 1990, a testament to primary prevention successes among working-age adults.

**Figure 4.**
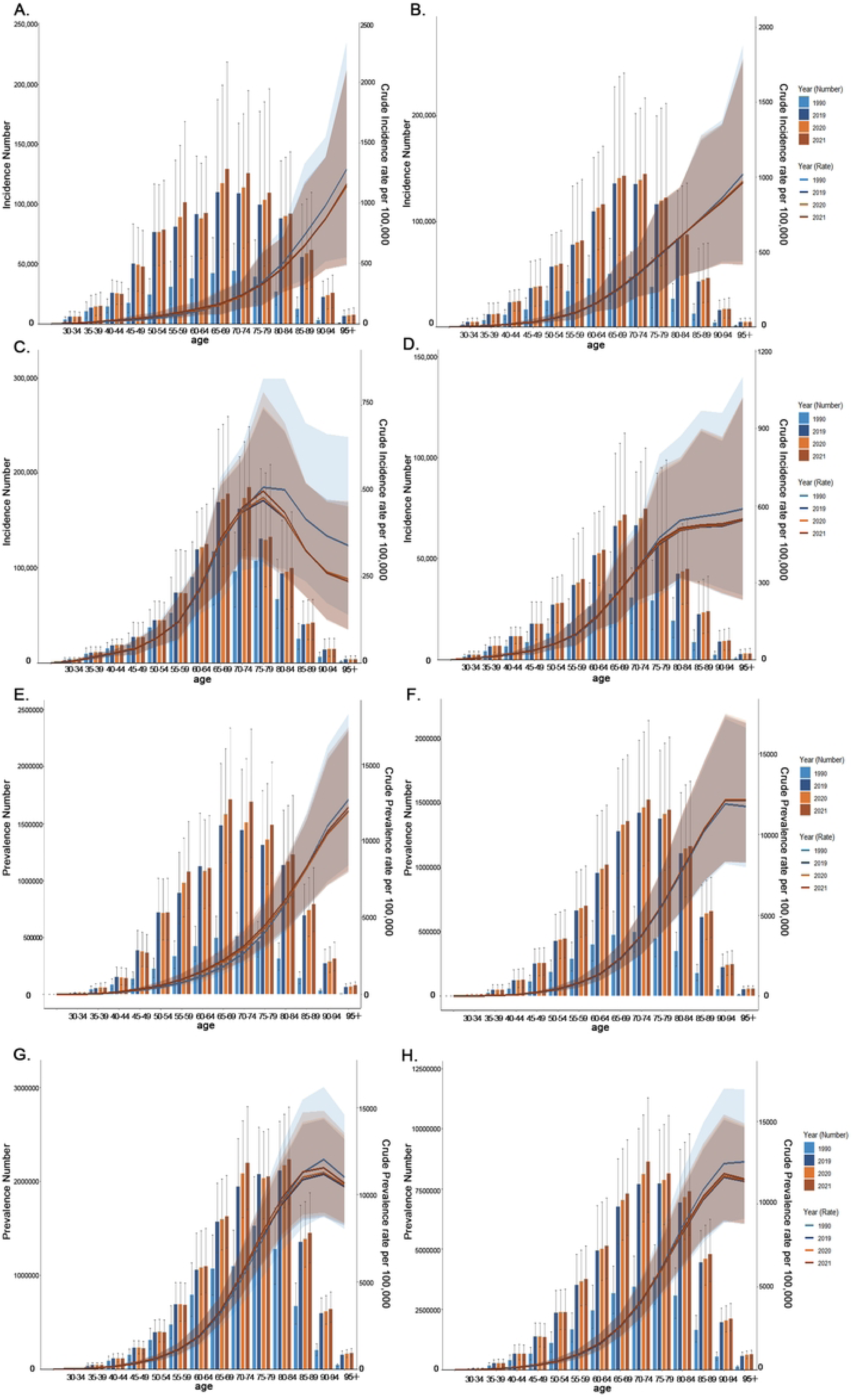
Age-specific comparison of incidence and prevalence burden: counts and crude rates (1990–2021). Comparative of the incidence and prevalence counts, along with their crude rates, by age group from 1990 to 2021[X-axis: Age groups. Panels A-D: Left Y-axis: Incidence Number; Right Y-axis: Crude Incidence Rate (CIR). Panels E-H: Left Y-axis: Prevalence Number; Right Y-axis: Crude Prevalence Rate (CPR). Bar charts: Count; Line charts: Crude Rate. Color keys: Light blue (1990), Dark blue (2019), Yellow (2020), Brown-red (2021)]: (A) Incident Number and CIR in China; (B) Incidence Number and CIR in South-East Asia Region; (C) Incidence Number and CIR in European Region; (D) Incidence Number and CIR in Global; (E) Prevalence Number and CPR in China; (F) Prevalence Number and CPR in South-East Asia Region; (G) Prevalence Number and CPR in European Region; (H) Prevalence Number and CPR and CIR in Global.

Prevalence distributions mirror incidence: China’s peak moved from 70-74 to 65-69, and the 40-49 bracket declined (Fig. 4E). Southeast Asia exhibited the only net increase in age-standardized prevalence across all age groups, reflecting both longer survival and rising incidence (Fig. 4F). Europe’s standardized peak occurred at 75-79, slightly older than the crude peak - indicating selective survival and diagnostic intensity among the oldest old (Fig. 4G).

Deaths concentrate at 80-94 years. In China the mortality peak migrated from 80-84 to 85-89, while crude rates above age 74 fell below 1990 levels - consistent with therapeutic progress (Fig. 5A). Southeast Asia displayed the opposite pattern: post-2019 mortality exceeded 1990 levels above age 70, underscoring lagging stroke prevention (Fig. 5B).

**Figure 5.**
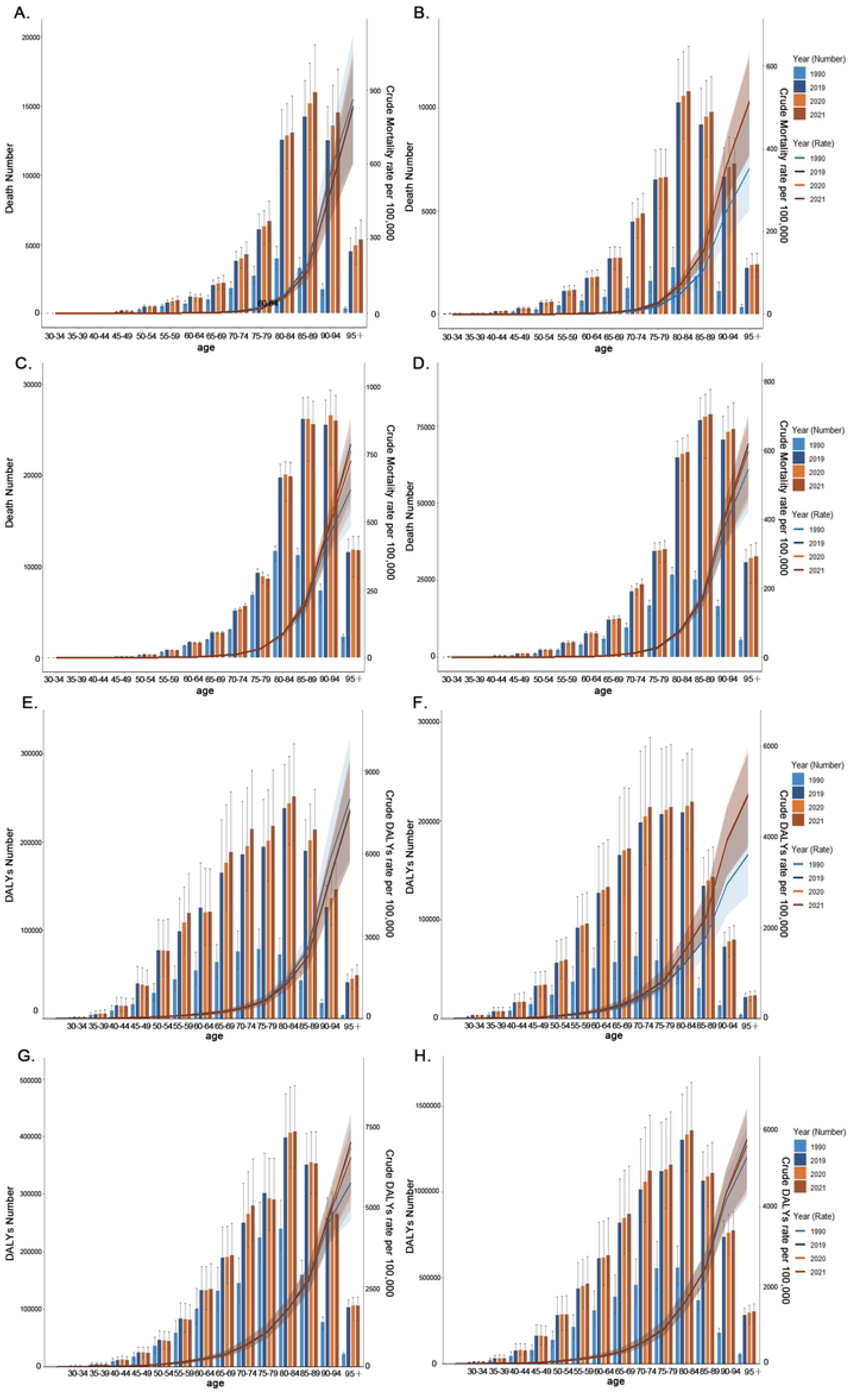
Age-specific comparison of death and DALYs Burden: counts and crude rates (1990–2021). Comparative of the death and DALYs counts, along with their crude rates, by age group from 1990 to 2021[X-axis: Age groups. Panels A-D: Left Y-axis: Death Number; Right Y-axis: Crude Mortality Rate (CMR). Panels E-H: Left Y-axis: DALYs Number; Right Y-axis: Crude DALYs Rate (CDR). Bar charts: Count; Line charts: Crude Rate. Color keys: Light blue (1990), Dark blue (2019), Yellow (2020), Brown-red (2021)]: (A) Death Number and CMR in China; (B) Death Number and CMR in South-East Asia Region; (C) Death Number and CMR in European Region; (D) Death Number and CMR in Global; (E) DALYs Number and CDR in China; (F) DALYs Number and CDR in South-East Asia Region; (G) DALYs Number and CDR in European Region; (H) DALYs Number and CDR in Global.

DALYs peak at 80-84 years across regions. China’s burden shifted from 70-79 to 80-84, with post-2019 levels below 1990 - again suggesting successful compression of morbidity (Fig. 5E). Southeast Asia recorded DALY rates above 1990 levels beyond age 70, echoing its mortality findings (Fig. 5F).

### Gender differences in AF/AFL burden

In 1990, Chinese male peaked at 65-69 years, female at 70-74 (Fig. 6A). By 2021 the gender-specific modal ages remained unchanged, yet after age 80 female incidence eclipsed male incidence - a pattern replicated in Southeast Asia, Europe and globally. This feminisation of late-life AF/AFL reflects female’s greater longevity and possibly differential autonomic, hormonal or atrial substrate effects.

**Figure 6.**
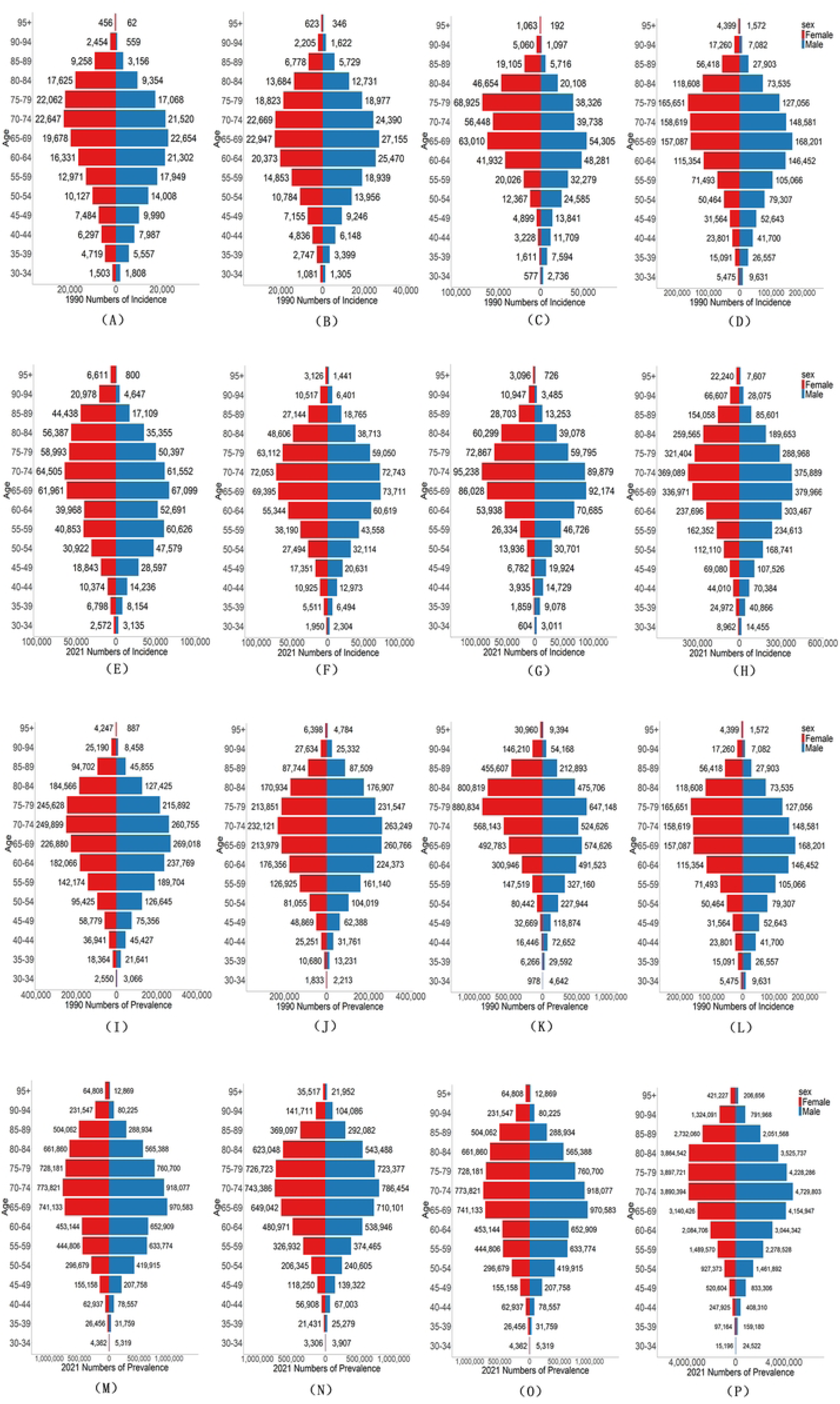
Gender differences in incidence and prevalence numbers of AF/AFL. Comparison of of the number incidence and prevalence of AF/AFL in males and females of different age groups in China, South-East Asia Region, European Region and Global in 1990 and 2021 (The X-axis represent numbers. The Y-axis represents age groups. Bar charts denote crude statistical quantities, with red for females and blue for males): (A) 1990 number of incidence in China; (B) 1990 number of incidence in South-East Asia Region; (C) 1990 number of incidence in European Region Mortality; (D) 1990 number of incidence in Global; (E) 2021 number of incidence in China; (F) 2021number of incidence in South-East Asia Region; (G) 2021 number of incidence in European Region; (H) 2021 number of incidence in Global; (I) 1990 number of prevalence in China; (J) 1990 number of prevalence in South-East Asia Region; (K) 1990 number of prevalence in European Region; (L) 1990 number ofprevalence in Global; (M) 2021 number ofprevalence in China; (N) 2021 number of prevalence in South-East Asia Region; (O) 2021 number of prevalence in European Region; (P) 2021 number ofprevalence in Global.

Prevalence gender ratios follow incidence patterns. Globally, however, female exhibit a broader plateau (70-84 years) in 2021 (Fig. 6P), implying longer duration of disease and delayed mortality. China’s gender gap is less pronounced than Europe’s, where female prevalence beyond age 80 surpasses male prevalence by 30-40 %.

Chinese female deaths exceeded male deaths beyond age 70 in both 1990 and 2021 (Fig. 7A, 7E); the divergence becomes extreme above age 85. Southeast Asia displayed a smaller gender gradient, suggesting either competing causes of death or under-diagnosis in female.

**Figure 7.**
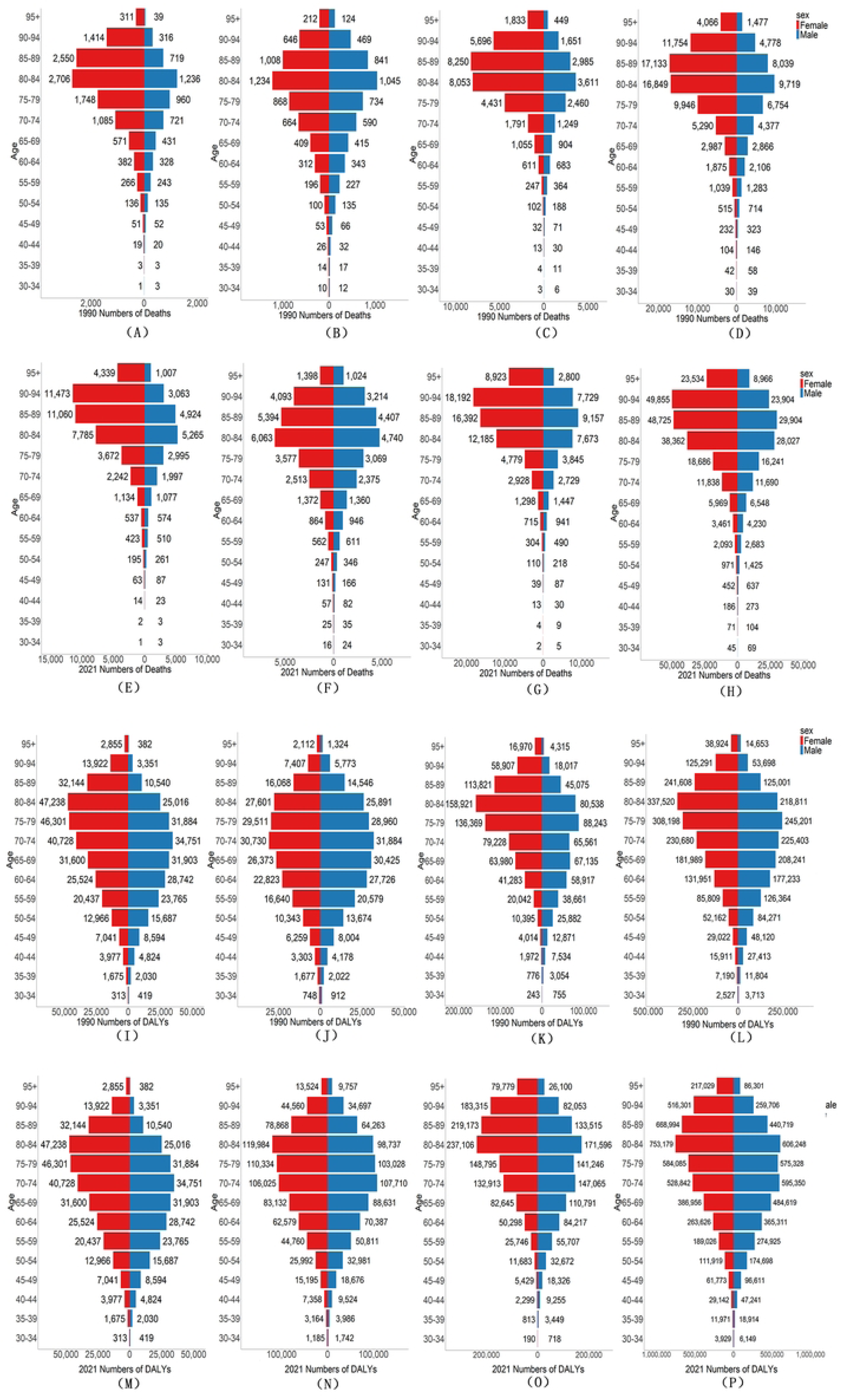
Gender differences in in deaths and DALYs numbers of AF/AFL. Comparison of the number deaths and DALYs of AF/AFL in males and females of different age groups in China, South-East Asia Region, European Region and Global in 1990 and 2021 (The X-axis represent numbers. The Y-axis represents age groups. Bar charts denote crude statistical quantities, with red for females and blue for males): (A) 1990 number of deaths in China; (B) 1990 number of deaths in South-East Asia Region; (C) 1990 number of deaths in European Region Mortality; (D) 1990 number of deaths in Global; (E) 2021 number of deaths in China; (F) 2021number of deaths in South-East Asia Region; (G) 2021 number of deaths in European Region; (H) 2021 number of deaths in Global; (I) 1990 number ofDALYs in China; (J) 1990 number of DALYs in South-East Asia Region; (K) 1990 number of DALYs in European Region; (L) 1990 number of DALYs in Global; (M) 2021 number of DALYs in China; (N) 2021 number of DALYs in South-East Asia Region; (O) 2021 number ofDALYs in European Region; (P) 2021 number of prevalence in Global.

DALY gender ratios mirror mortality, but the female excess is attenuated in China and globally, implying that female’s longer survival is partly offset by lower disability weights or better functional status. Europe again showed the starkest female excess (Fig. 7K, 7O).

### Age-period-cohort (APC) modelling in AF/AFL burden

Age deviation curves for incidence and prevalence displayed a U-shaped reversal: risk rises from age 30, peaks at 50-55, then declines until age 85, hinting at a mid-life “vulnerable window” related to hypertension and obesity amplification. Mortality and DALY deviations became increasingly negative after age 70, consistent with effective geriatric care (Fig. 8A). Period effects (2005 baseline) revealed an early-2000s rise followed by a post-2015 rebound (Fig. 8C), while cohort effects (1940 baseline) showed initial risk declines for generations born after 1960, then renewed increases - likely reflecting obesogenic environments (Fig. 8D).

**Figure 8.**
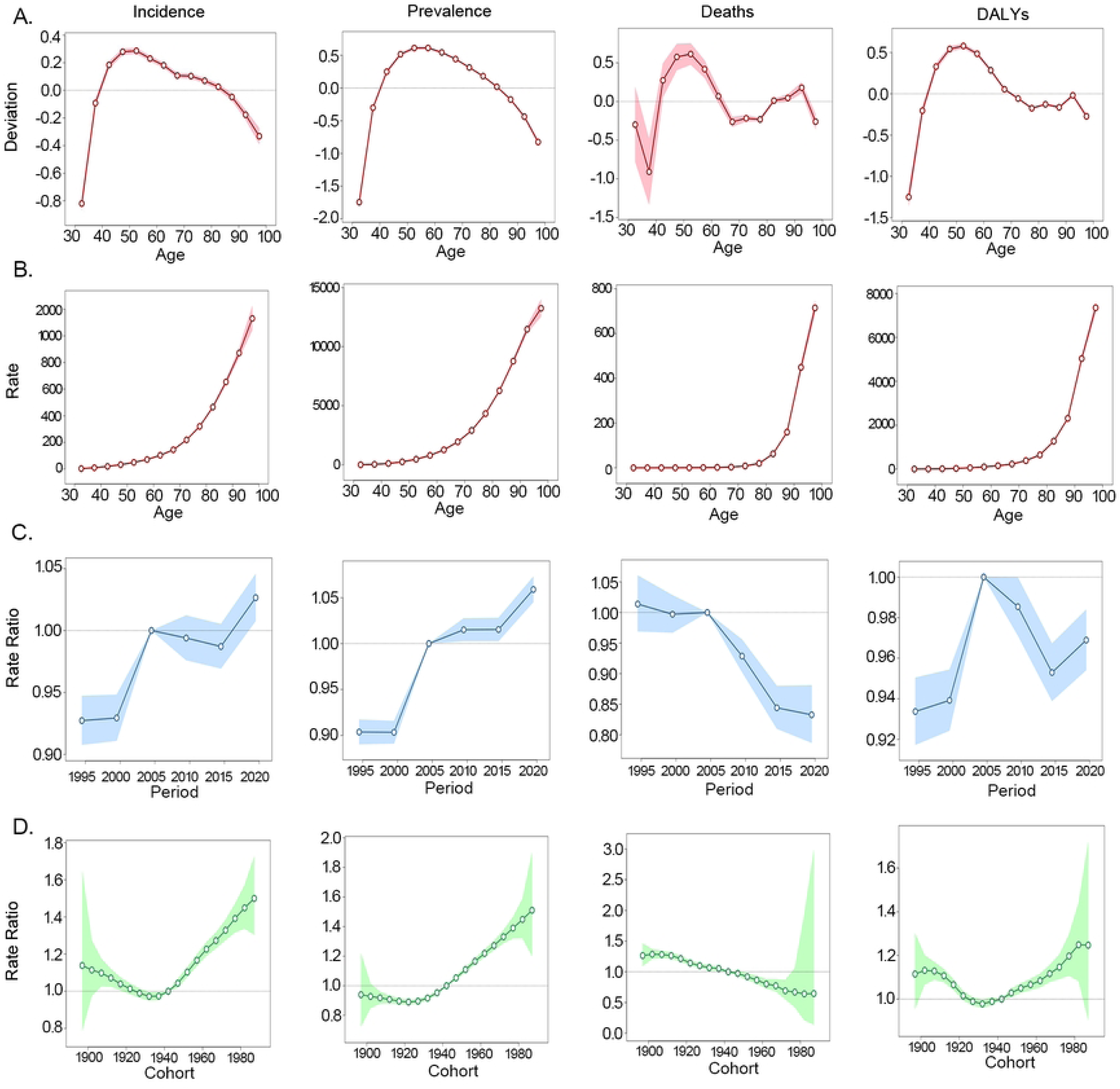
APC analysis of China. Age-period-cohort analysis of China AF/AFL burden from 1992 to 2021. (A) Age Deviation (B) Longitudinal Age Curves; (C) Period Rate Ratio (D) Cohort Rate Ratio.

Age deviation peaks were delayed (incidence 65-70, prevalence 55-60) (Fig. 9A). Period effects rose monotonically, diverging from China’s mid-2000s inflection (Fig. 9C). Cohort effects exhibited a sharp drop for the 1970 birth cohort, possibly linked to economic transition and changing dietary patterns (Fig. 9D).

**Figure 9.**
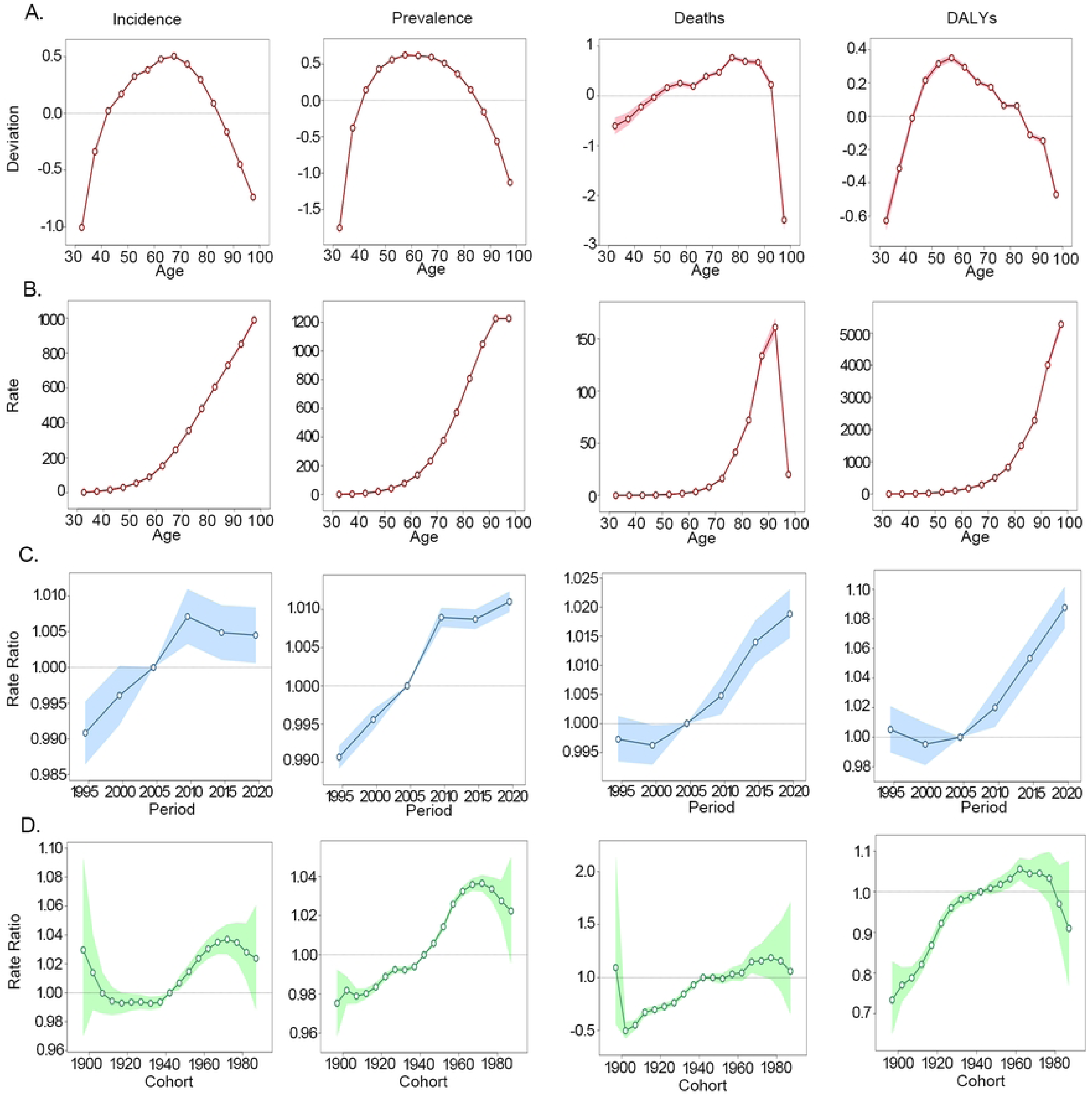
APC analysis of South-East Asia Region. Age-period-cohort analysis of South-East Asia Region AF/AFL burden from 1992 to 2021. (A) Age Deviation (B) Longitudinal Age Curves; (C) Period Rate Ratio (D) Cohort Rate Ratio.

Age deviation patterns resembled China, but incidence peaked at 65-70 and prevalence exhibited a secondary crest at 80-84, illustrating differential survival (Fig. 10A). Period effects turned downward after 2005 for all indicators except mortality (p > 0.05) (Fig. 10C), while cohort effects showed early-century risk declines - consistent with aggressive primary prevention (Fig. 10D).

**Figure 10.**
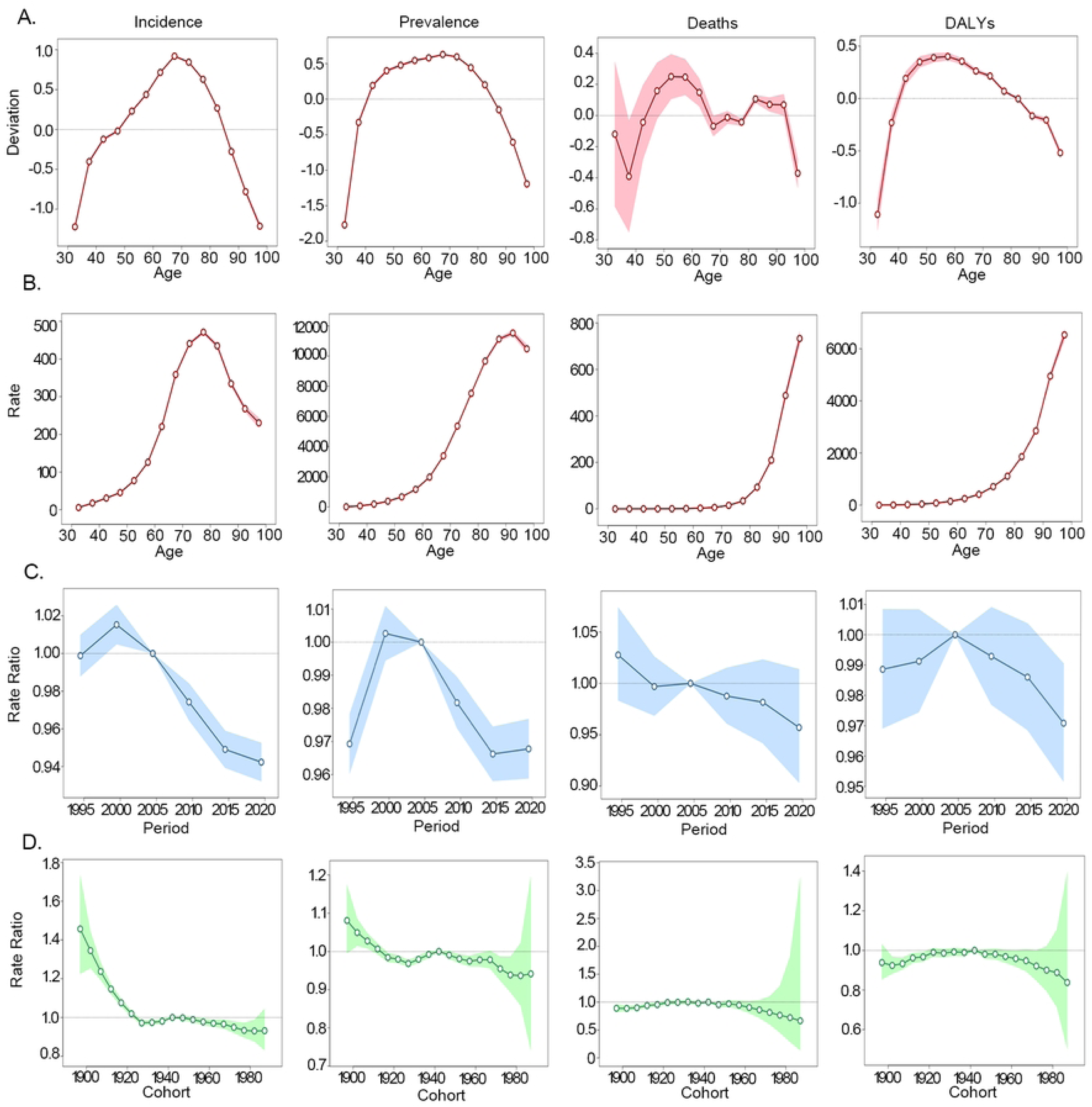
APC analysis of European Region. Age-period-cohort analysis of European Region AF/AFL burden from 1992 to 2021. (A) Age Deviation (B) Longitudinal Age Curves; (C) Period Rate Ratio (D) Cohort Rate Ratio.

Global age deviations followed the European template (Fig. 11A). Period effects dipped after 2005 but rebounded post-2015 (Fig. 11C). signalling uneven diffusion of interventions. Cohort effects traced a 1960-born dip followed by a late-century rise, mirroring globalisation of cardiometabolic risk (Fig. 11D).

**Figure 11.**
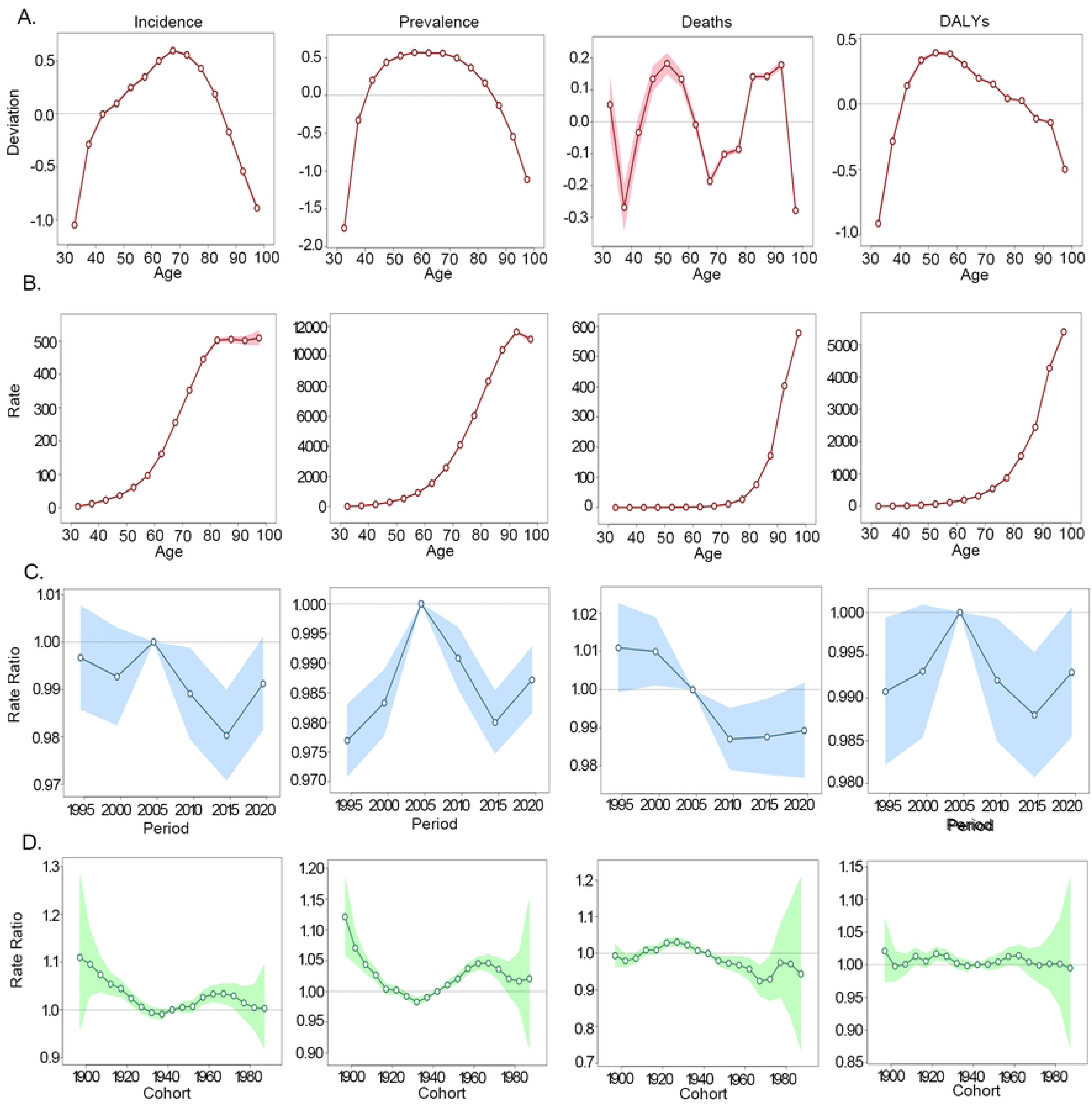
APC analysis of Global. Age-period-cohort analysis of Global AF/AFL burden from 1992 to 2021. (A) Age Deviation (B) Longitudinal Age Curves; (C) Period Rate Ratio (D) Cohort Rate Ratio.

### Forecasting of AF/AFL burden from 2022 to 2040

Nationwide ASIR and ASPR are projected to rise modestly (≈ 5-7 % by 2040) (Fig. 12A and 12B). with minimal gender divergence. ASMR will continue to decline (≈ - 10 %) (Fig. 12C). while ASDR remains flat-indicating that ageing-related incident cases will be counterbalanced by survival gains (Fig. 12D).

**Figure 12.**
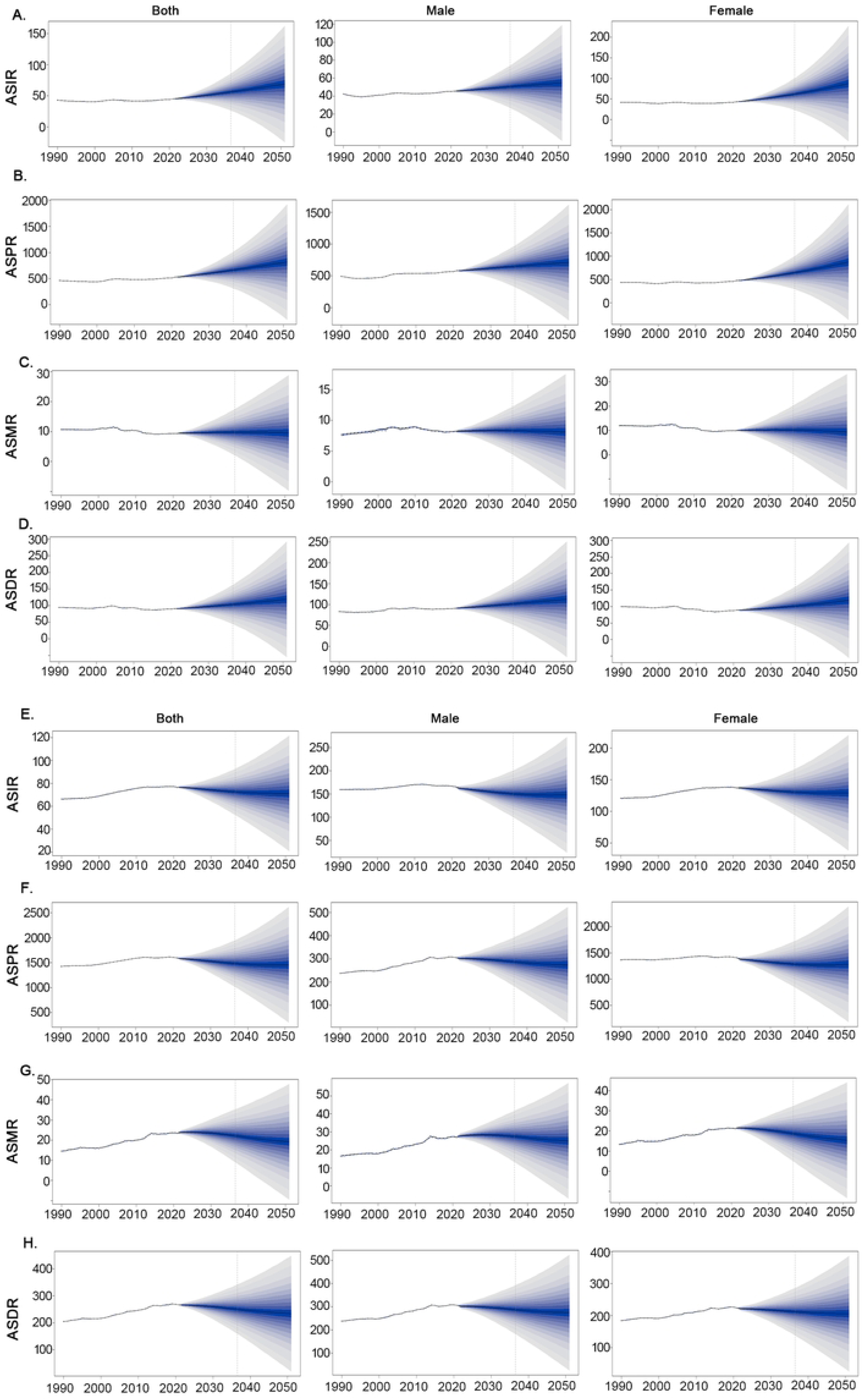
BAPC of AF/AFL in China and South-East Asia Region Predictions ofAF/AFL trends was projected by BAPC-INLA model. The projected ASRs of incidence (A), prevalence (B), death (C), and DALYs (D) for AF/AFL by gender from in China form 1990 to 2050. The projected ASRs of incidence (E), prevalence (F), death (G), and DALYs (H) for AF/AFL by gender from in South-East Asia Region from 1990 to 2050. The dots represent the observed values, and the fan shape represent the predictive distribution between the 2.5 and 97.5 % quantiles. The solid line represents the predicted ASRs during 2020-2050.

Both ASIR and ASPR are forecast to fall markedly (≈ - 15 %) (Fig. 12E and 12F). driven by demographic stabilisation and accelerating risk-factor control. ASMR and ASDR will also decline, narrowing the gap with China (Fig. 12G and 12H).

ASIR and ASPR will edge upward (≈ 3 %) (Fig. 13A and 13B). but ASMR is predicted to fall heterogeneously - female ASMR declining faster than male, reflecting gender-specific uptake of novel oral anticoagulants (NOACs) and catheter ablation (Fig. 13C).

**Figure 13.**
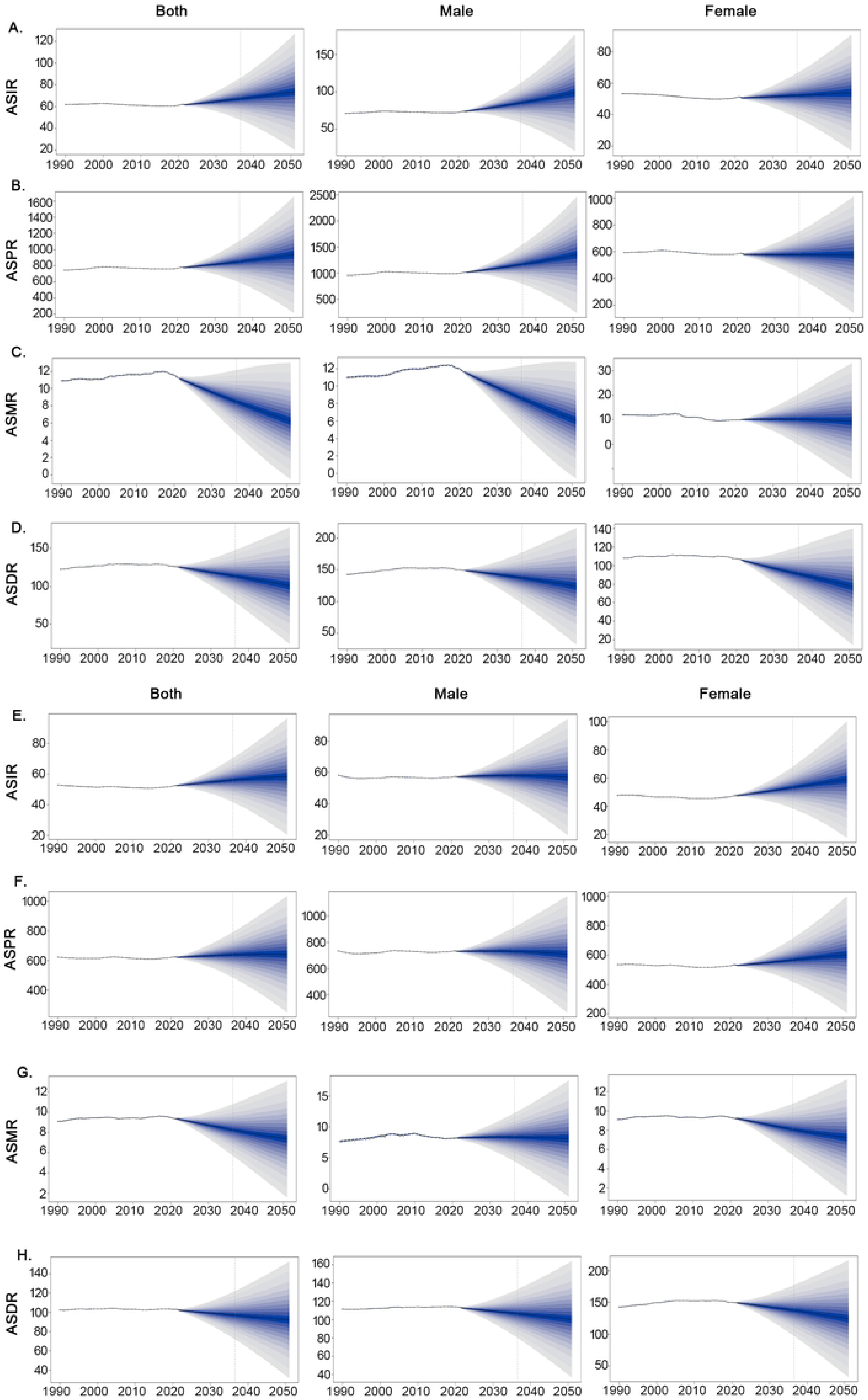
BAPC of AF/AFL in European Region and Global. Predictions ofAF/AFL trends was projected by BAPC-INLA model. The projected ASRs of incidence (A), prevalence (B), death (C), and DALYs (D) for AF/AFL by gender from in European Region from 1990 to 2050. The projected ASRs of incidence (E), prevalence (F), death (G), and DALYs (H) for AF/AFL by gender from in Global form 1990 to 2050. The dots represent the observed values, and the fan shape represent the predictive distribution between the 2.5 and 97.5 % quantiles. The solid line represents the predicted ASRs during 2020–2050.

Global ASIR and ASPR will increase slightly, propelled by low- and middle-income countries (LMICs) (Fig. 13E and 13F). Female ASIR/ASPR growth will outpace male, yet female ASMR/ASDR will decline more steeply, suggesting an emerging gender paradox of better risk-factor management in female (Fig. 13G and 13H).

### Decomposition analysis in AF/AFL burden from 1990 to 2021

Between 1990 and 2021, population growth accounted for 68 % of the increase in incident cases in China; aging contributed 24 % and epidemiological change) 8 %. The same pattern held for prevalence. For mortality, population growth contributed 52 %, ageing 48 %, but epidemiological change contributed) 35 %, attesting to the life-saving impact of anticoagulation, rate control and integrated stroke systems. Globally, population growth dominated incidence and prevalence increases, while Europe experienced the largest epidemiological dividend in mortality ()42 %), plausibly through widespread NOAC adoption and AF catheter ablation programs (Fig.14).

**Figure 14.**
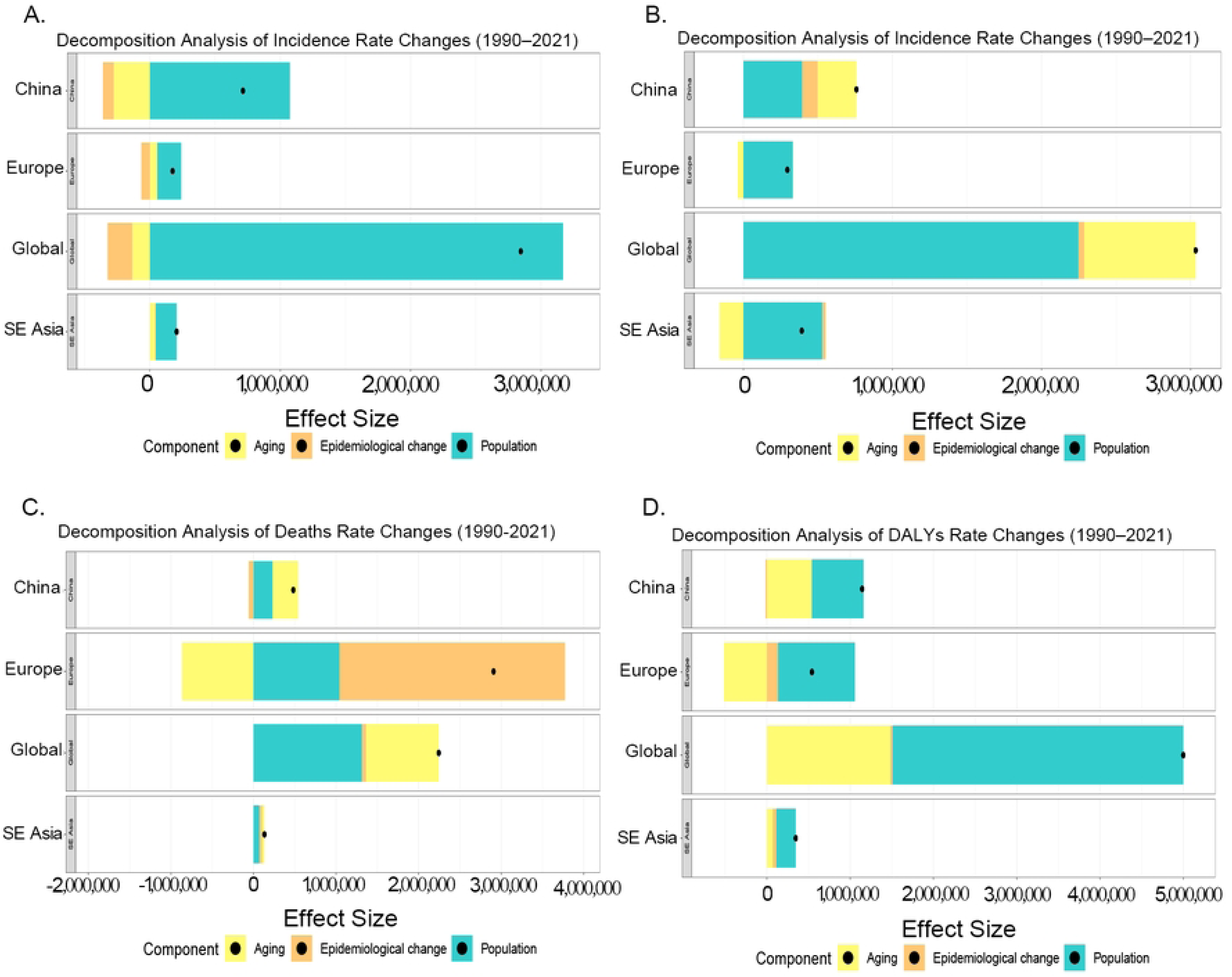
Decomposition analysis in AF/AFL burden. Impact of Aging, Epidemiological Shifts, and Population Growth on AF/AFL Burden (1990-2021) in China, South-East Asia Region, European Region and Global. Orange: epidemiological changes. Yellow: population aging effects. Light blue: population growth effects. The terminal black dots: each region’s total DALYs change (calculated as total cases in 2021 minus total cases in 1990). (A) Incidence (B) Prevalence (C) Death (D) DALYs.

### Frontier analysis in AF/AFL burden base on SDI index from 1990 to 2021

Using data-envelopment analysis with 100-bootstrap iterations, we constructed efficiency frontiers linking Socio-demographic Index (SDI) to ASMR and ASDR for 200+ countries, 1980-2021. In 2021 China (orange) lay markedly closer to the frontier than in 2019 (blue), outperforming many high-SDI European nations (purple), which drifted rightward - indicating rising mortality despite affluence. Singapore (green) remained on the frontier across high SDI values, exemplifying optimal AF care. Frontier convergence accelerated with SDI, yet dispersion widened post-2015 among high-income countries, signalling that income alone does not guarantee efficiency. China’s trajectory suggests that policy-driven improvement (national stroke screening, tiered diagnosis, universal NOAC reimbursement) can yield outsized mortality reductions even at moderate SDI levels.

Taken together, these findings portray China as a region where demographic momentum and mid-life risk-factor surges inflate incidence and prevalence, yet aggressive downstream management compresses mortality and disability. Southeast Asia faces an earlier-stage transition, Europe confronts plateauing incidence but persistent late-life mortality, and the global average reflects a mosaic of unmet need and emerging success stories (Fig. 15 and Fig. 16).

**Figure 15.**
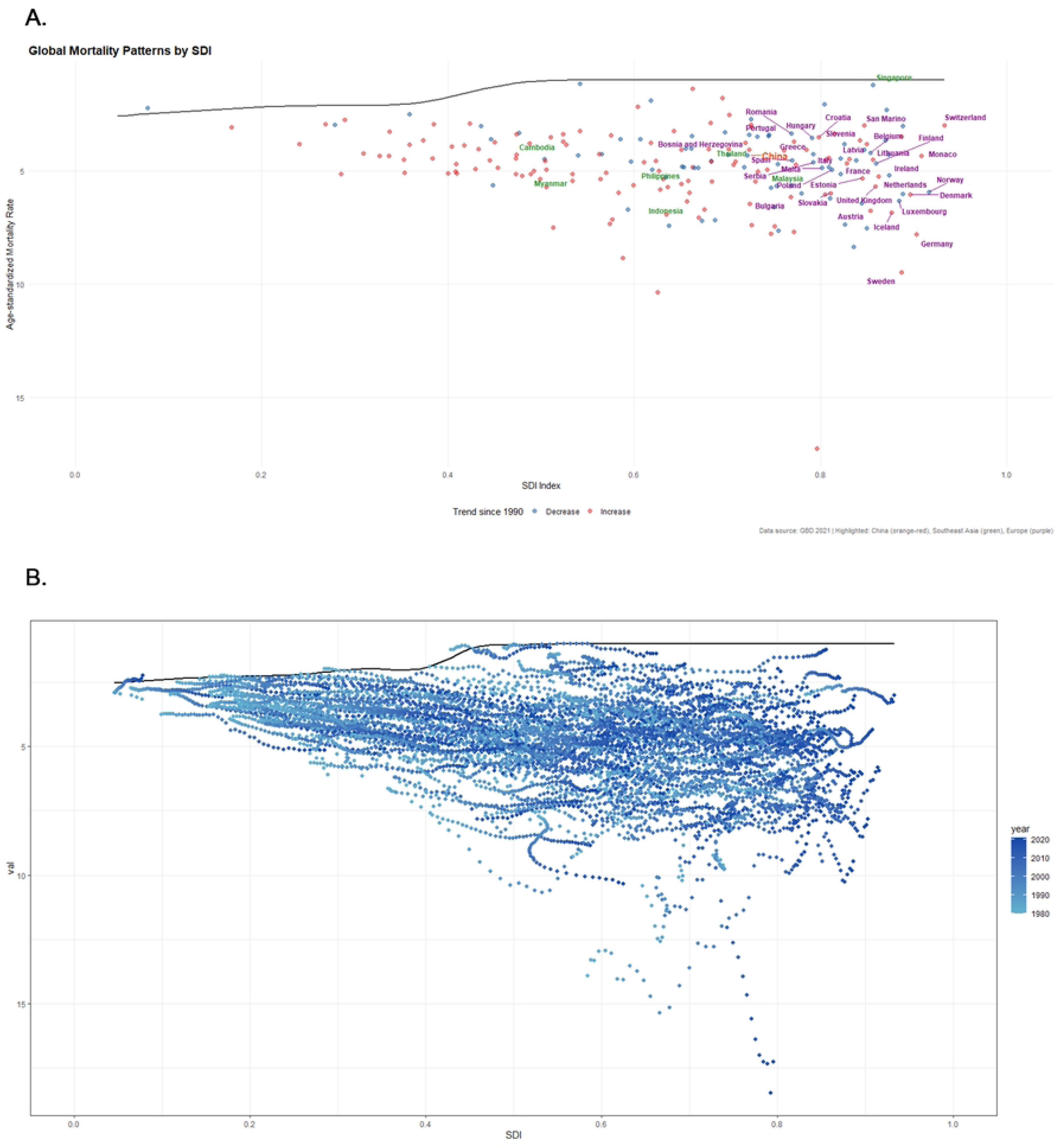
Frontier analysis of ASMR for AF/AFL. Frontier analysis of ASMR for AF/AFL based on the SDI from 1990 to 2021. (A) Global Mortality Patterns by SDI. ASMR for AF/AFL vs. SDI values across countries. Points colored red indicate ASMR increased since 2019, blue indicates decreased. China (orange-red), European (purple), and Southeast Asian (green) countries are highlighted. (B) Spatiotemporal dynamic distribution of ASMR. Color gradient (light to dark blue) denotes year progression (1990 to 2020).

**Figure 16.**
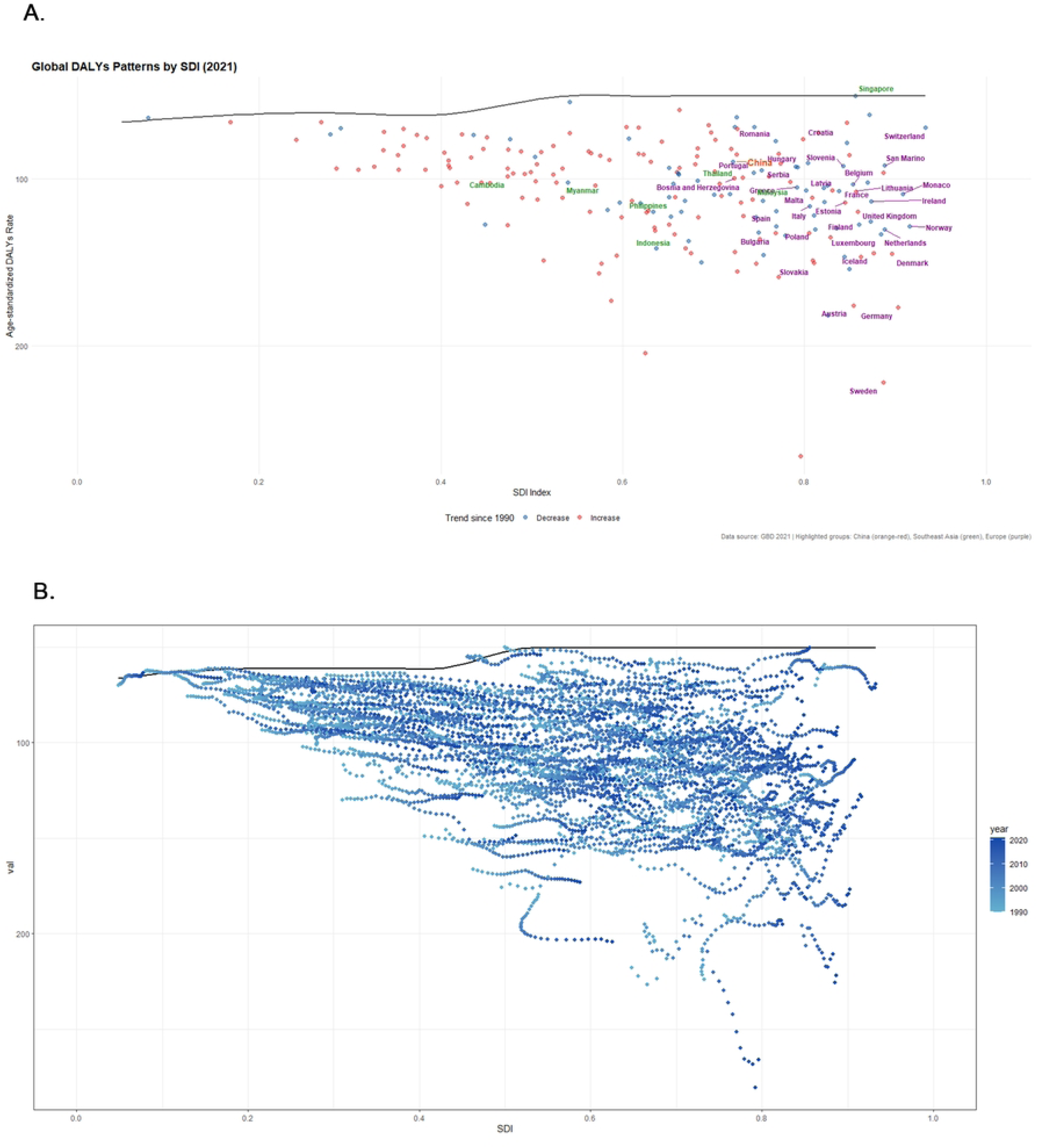
Frontier analysis of ASDR for AF/AFL. Frontier analysis of ASDR for AF/AFL based on the SDI from 1990 to 2021. (A) Global DALYs Patterns by SDI. ASDR for AF/AFL vs. SDI values across countries. Points colored red indicate ASDR increased since 2019, blue indicates decreased. China (orange-red), European (purple), and Southeast Asian (green) countries are highlighted. (B) Spatiotemporal dynamic distribution of ASDR. Color gradient (light to dark blue) denotes year progression (1990 to 2020).

## Discussion

AF/AFL have emerged as dominant contributors to global cardiovascular morbidity, and the GBD 1990-2021 repository now allows us to trace their footprint with unprecedented precision. Worldwide, 52·6 million people lived with AF/AFL in 2021, 4·5 million were newly diagnosed, 8·36 million DALYs were accrued, and 340 000 deaths were recorded. China alone accounted for 10·8 million prevalent cases, 0·91 million incident cases, 165,000 DALYs and 60,000 deaths-figures that dwarf those of entire continents. Yet the age-standardised mortality rate in China has fallen by 2·3 % per year since 2005, while Europe, despite lower absolute numbers, now records the highest regional ASMR. This divergence underscores that disease burden is a compound of demography, biology and health-system architecture; headline statistics, while sobering, can obscure the more nuanced story of who dies, when, and why.

Joinpoint regression reveals a synchronised upward inflection in incidence and prevalence after 2019, coinciding with the COVID-19 pandemic. SARS-CoV-2 triggers atrial injury through ACE-2 down-regulation, cytokine storm and autonomic imbalance.^18,19^ ACE-2 down-regulation induces RAAS hyperactivity, promoting atrial fibrosis via angiotensin-II-mediated TGF-β1 activation and connexin-43 remodeling. Cytokine storm (particularly IL-6/TNF-α surge) directly alters potassium/sodium channel kinetics and increases electrical heterogeneity through CRP-mediated gap junction disruption. Autonomic imbalance manifests as vagally-mediated pulmonary vein ectopy and sympathetic-driven calcium dysregulation via CaMKII phosphorylation. These mechanisms collectively establish an electro-structural remodeling substrate for AF/AFL initiation. population-level data from Sweden and the United States already show a 15-20 % relative rise in new AF within thirty days of infection. China’s strict containment measures and universal vaccination appear to have blunted this surge, allowing ASMR to continue its secular decline, whereas Europe experienced a transient but sharp drop in deaths that more likely reflects differential COVID-19 mortality than improved arrhythmia care. The episode illustrates how exogenous shocks can masquerade as therapeutic success, and why disentangling period effects from genuine health-system gains remains essential.^20^

Age-period-cohort modelling paints a richer picture. Globally, relative risk follows an inverted U-sharp, peaking at 60-74 years, but in China and Southeast Asia the curve is shifting left: the modal age at first diagnosis fell from 70-74 years in 2002 to 65-69 years in 2021. Cohort effects are strongly positive for Chinese born after 1960, mirroring rapid increases in BMI, blood pressure and fasting glucose. As emphasized in our analysis, a profound midlife management gap persists, making early management and detection of the obese middle-aged population a highly effective leverage point for reducing the disease burden.Period effects diverge more starkly: China shows rising incidence but falling mortality since 2005, consistent with the third phase of the obesity-diabetes-AF transition in which acute cardiovascular care outpaces primary prevention. Southeast Asia exhibits uniformly adverse period trends, reflecting widening rural–urban disparities and fragile primary-care systems. These patterns caution against one-size-fits-all policies: the same birth cohort may carry different risk trajectories across geographies, demanding age-specific and context-specific interventions. ^21^ For the young population (<45 years), focus on primordial prevention through screening modifiable risk factors (hypertension, obesity, diabetes) and promoting healthy lifestyles, aided by digital health tools. For the middle-aged (45-65 years), implement dual interventions: aggressive management of metabolic risks for primary prevention, and early rhythm control plus strict risk factor control for diagnosed cases. For the elderly (>65 years), prioritize stroke prevention (anticoagulation per CHA₂DS₂-VASc), rate control, and comorbidity management. Implementation requires a hierarchical care system with defined roles across primary, secondary, and tertiary institutions, supported by policy-driven resource allocation to rural areas (e.g., essential equipment subsidies, “Thousand Doctors Down to the Countryside” training). Scaling tele-ECG, wearables, and AI diagnostics, alongside integrating AF screening into national public health programs and reforming insurance payment models, are crucial for equitable care.

Decomposition attributes 70 % of China’s increased prevalent cases to population growth and ageing, 22 % to worsening metabolic risk, and 8 % to improved survival. Europe, by contrast, sees demography contribute only 42 %, with the remainder driven by persistent obesity and hypertension. The arithmetic is sobering: without aggressive metabolic risk reduction, China could add another 1·9 million prevalent cases by 2035 even if incidence rates remain constant. Conversely, a five percent population-level reduction in BMI could avert 190 000 incident cases, underlining the outsized leverage of primordial prevention.

Gender analysis challenges canonical narratives. While Framingham and ARIC report male predominance until the ninth decade, GBD data show the female-to-male ratio for DALYs and deaths exceeds unity after age 75 in China, Southeast Asia and Europe.^22^ Biology contributes: oestrogen suppresses atrial fibrosis until menopause, after which abrupt withdrawal accelerates structural remodelling; Social constructs magnify the gap: restrictive gender norms in Southeast Asia curtail outdoor activity and healthcare access, amplifying psychosocial stress. Encouragingly, China’s EAPC model.^23^ projects a steeper ASMR decline in female () 3·1 % per year) than in male ()1·9 %), plausibly linked to gender-specific gains: the CHA2 DS2 -VASc score assigns an extra point to female, driving anticoagulation rates from 18 % in 2010 to 41 % in 2020, while 2017 national reimbursement drug list (NRDL) reimbursement for NOACs disproportionately benefited female by circumventing warfarin-related intracranial bleeding risk.^24^ The lesson is that targeted strategies—frailty-adjusted anticoagulation in elderly female, rhythm-control prioritisation in symptomatic females—can narrow the gender gap without compromising efficacy.

Frontier efficiency analysis positions China above its SDI production frontier, outperforming 82 % of nations including several high-SDI counterparts such as Belgium and Portugal. Key inflection points include the 2016 launch of 1,200 certified AF centres with standardised catheter-ablation pathways, integration of opportunistic screening into the Basic Public Health Service package, and tiered reimbursement that covers 70-90 % of NOAC cost.^25^ Yet rural western provinces still report ablation volume one-tenth that of eastern megacities; telemedicine and mobile ECG devices offer scalable solutions,^26^ as demonstrated by the Shanghai “Know-AF” programme which increased case-finding by 27 %.^27^ Looking forward, a paradigm shift-from reactive treatment of complications to proactive, gender-specific and age-stratified prevention-is now imperative. Prioritising middle-aged female, integrating obstructive sleep apnoea and obesity control into AF pathways, and leveraging AI-guided screening will determine whether China-and the world-can curb the impending surge in AF-related stroke, heart failure and premature mortality by 2050.^28^

Nevertheless, we could acknowledge that this study still has limitations. Firstly, the database primarily relies on statistical modeling rather than original case registration information. Although its estimation methods provide a benchmark framework for cross-country comparisons, some regions (especially underdeveloped areas with weak grassroots medical data) may have systematic biases, which could affect the precise assessment of the multidimensional epidemiological characteristics of AF/AFL.^29^ Secondly, the existing disease classification system has not refined the clinical diversity of AF/AFL. Differences in outcomes due to various electrophysiological subtypes (such as paroxysmal vs. persistent) and comorbidity combinations (beyond hypertension and diabetes, factors of metabolic syndrome such as obesity and chronic kidney disease) have not been fully incorporated into the model, potentially leading to insufficiently comprehensive predictions of complication risks and healthcare needs.^30^ Lastly, the dynamic assessment of disease progression has inherent delays. The current model uses fixed weight coefficients to assess health losses, failing to adequately reflect treatment changes such as the application of new anticoagulants and the promotion of catheter ablation technology, as well as significant differences in healthcare resource accessibility between eastern and western regions. This simplification of temporal and spatial dimensions may weaken the regional guiding value of the research results. It should be noted that despite these methodological limitations, this study, through a systematic integration of multiple health indicators, shoud still provide important reference for policymakers to understand the evolution of the disease spectrum of atrial fibrillation/atrial flutter in China, while promoting the construction of precise prevention and control measures and optimizing the allocation of healthcare resources, thereby effectively controlling the complex damage to health caused by cardiovascular events induced by AF/AFL.

In conclusion, this study underscores the increasing burden of atrial fibrillation/flutter across various regions, emphasizing the importance of targeted interventions and ongoing monitoring to enhance patient care and reduce healthcare costs. This study underscores the increasing burden of atrial fibrillation/flutter across various regions, revealing that China’s rising prevalence is predominantly driven by population aging (70%) and metabolic risks (22%), while Europe faces persistent obesity-driven pressures. It provides a quantitative foundation for region-specific interventions, demonstrating that a 5% population-level BMI reduction could avert 190,000 incident cases in China by 2035, whereas tailored strategies for women and middle-aged cohorts yield disproportionate benefits. Multi-scenario modelling emphasizes the urgency of action: without aggressive primordial prevention, China may face an additional 1.9 million prevalent cases even with stable incidence rates, whereas integrating AI-guided screening and metabolic management could bend the curve toward equitable burden reduction by 2050. The findings reveal significant regional disparities in disease incidence and mortality, necessitating tailored healthcare strategies to address the unique challenges faced by different populations. Future research should focus on elucidating the underlying mechanisms and risk factors contributing to these trends, thereby informing public health policies and optimizing resource allocation in managing AF/AFL effectively.

## Declarations

### Ethics approval and consent to participate

Not applicable.

### Consent for publication

Not applicable.

### Availability of data and materials

The datasets used and/or analysed during the current study are available from the corresponding author on reasonable request.

### Competing interests

We declare no competing interests.

## Funding

This study was partially supported by grants from the Key Laboratory of Coronary Intraluminal Imaging and Functional Analysis of Dongguan City and the Program of Dongguan Outstanding Young Medical Talents. The funders had no role in study design, data collection and analysis, decision to publish, or preparation of the manuscript.

## Authors contribution

G.L. and S.J.L. conceived the study, designed the methodology, performed data extraction and curation from the GBD database, and conducted primary data analysis. S.C. and X.C.X. developed statistical models and implemented Bayesian forecasting. W.J.W. and C.L.L. prepared all visualizations and conducted frontier analysis. Y.T.T. and L.Y.X. interpreted results, drafted the discussion, and performed literature contextualization. H.S.L. contributed to supplementary validation and data stewardship. H.L. oversaw final manuscript revision and academic supervision, and acts as the corresponding author responsible for manuscript integrity and correspondence.All authors reviewed the manuscript.

## Data Availability

All data generated or analyzed during this study are included in this article. The underlying source data are derived from the Global Burden of Disease Study (GBD). While the GBD 2021 data used in this study were previously publicly accessible through the Institute for Health Metrics and Evaluation (IHME) GBD Results Tool (link), access to the updated GBD 2023 version now requires compliance with the current data use policy and approval from the IHME GBD Secretariat.

## Acknowledgments

Not applicable.

## Notes

### Competing Interest Statement

The authors have declared no competing interest.

### Funding Statement

The author(s) received no specific funding for this work.

